# Identification of Blood miRNA Biomarkers in Systemic Tuberculosis through Metadata Analysis

**DOI:** 10.1101/2025.08.12.25333471

**Authors:** Swathi Chadalawada, SR. Rathinam, Bharanidharan Devarajan

**Affiliations:** Department of Microbiology and Bioinformatics, Aravind Medical Research Foundation, Madurai - 625020, Tamil Nadu, India; Biomedical Sciences, Madurai Kamaraj University, Madurai - 625021, India; Uvea services, Aravind Eye Hospital, Madurai, India

**Keywords:** Active tuberculosis, Latent tuberculosis, Metadata, Small-RNA sequencing, Transcriptome, *Mycobacterium tuberculosis*

## Abstract

Individual studies of miRNA analysis from small-RNA sequencing data can produce contradicting results. However, metadata analysis is used to overcome inconsistent findings between different studies. Thus, we aim to identify dysregulated miRNAs in systemic tuberculosis (TB) patients through metadata analysis using small-RNA sequencing data. 131 samples from seven different datasets were downloaded from the Sequence Read Archive (SRA) database. Among them, 45 were healthy controls, 47 with active TB, and 39 with Latent TB (LTB). First, we identified differentially expressed (DEs) miRNAs in active TB and LTB samples compared to controls. A total of 52 miRNAs were filtered based on their role in TB-specific, active TB, LTB-specific, and disease progression. These miRNAs may play an important role in TB disease progression from LTB to active TB. Subsequently, we performed gene enrichment and network analysis for both upregulated and downregulated miRNAs. From that, we selected eight miRNAs, hsa-miR-155-5p, hsa-miR-223-3p, hsa-miR-32-3p, hsa-miR-374a-3p, hsa-miR-374a-5p, hsa-miR-582-5p, hsa-miR-320d, and miR-122-5p served as biomarkers based on their role in TB pathogenesis through PI3K-Akt signaling pathway, TNF-signaling pathway, phagosome, and NOD-like signaling pathway. For the first time, we performed a metadata analysis with small-RNA sequencing data from publicly available datasets and identified miRNAs that could serve as biomarkers for systemic TB, which require further experimental confirmation.

## Introduction

Tuberculosis (TB) is a lethal infectious disease caused by *Mycobacterium tuberculosis* (*MTB*) that affects the lungs (Cohen et al., 2018). The clinical symptoms of the infected individuals were classified as latent TB (LTB) or active TB (Wei et al., 2020). Early diagnosis of TB is critical for the treatment and prevention of transmission from one person to another. The conventional methods, such as microbial culture, the tuberculin skin test (TST), and PCR-based molecular testing methods, are still insufficient for diagnosing TB infection (Ulger et al., 2022). The current TB blood test methods, such as interferon-gamma released assay (IGRA), improved the sensitivity and specificity to detect active TB, and LTB was 90.80% and 76.62%, respectively (Ai et al., 2018). Several studies showed differential gene expression of cytokines and chemokines in active TB patients (Domingo-Gonzalez et al., 2016, Yu et al., 2012). The significant expression of CLEC3B, SELL, ECM1, and IP10 protein markers was identified in active pulmonary TB from healthy donors or *MTB*-infected individuals (Ndiaye et al., 2022). The gene *PTPRC* may distinguish active TB from healthy individuals, and *ASUN* discriminates LTB from healthy individuals. However, *DHX29* could identify LTB individuals from active TB or healthy individuals (Lee et al., 2016). Comparing the groups with TB and without TB showed that *KIR2DS2* was associated with an increased risk of TB onset (de Sá et al., 2020). Significant higher expression of *NPC2* mRNA as a diagnostic host biomarker for active TB also reveals its potential to predict progression from LTB to active TB and response to anti-TB treatment (de Araujo et al., 2021). These studies were limited by the small sample size and lack of functional studies to identify candidate biomarkers in TB pathogenesis. However, these studies are not sufficient to distinguish between active TB, LTBI, and disease progression. Recent reports have suggested that serum-derived miRNAs are highly stable under harsh conditions and can be used as biomarkers for various infectious diseases (Chakrabarty et al., 2019).

MicroRNAs are small non-coding RNA molecules with 22nt length and have an important role in disease pathogenesis (Ratti et al., 2020). Previous studies showed that altered blood miRNAs in tuberculosis patients could serve as biomarkers (Behrouzi et al., 2019, Sabir et al., 2018). Zhang H et al., 2014 indicated that hsa-miR-196b and hsa-miR-376c served as potential serum markers in active TB (Zhang et al., 2014). In peripheral blood mononuclear cells (PBMC) of active TB patients, blood samples showed abundant expression of hsa-miR-27a target CANA2D3, leading to the downregulation of Ca^2+^ signaling, thus inhibiting autophagosome formation and promoting increased intracellular survival of *MTB* by manipulating the Ca^2+^-associated autophagy (Liu et al., 2018). Araujo LS et al., 2019 showed that three miRNAs, hsa-let-7a-5p, hsa-miR-589-5p, and hsa-miR-196b-5p as a highly sensitive (100%) classifier to distinguish TB from non-TB groups in peripheral blood (de Araujo et al., 2019). Hashimoto S et al., 2020 demonstrated that eight miRNAs, hsa-miR-122-5p, hsa-let-7i-5p, hsa-miR-148a-3p, hsa-miR-151a-3p, hsa-miR-21-5p, hsa-miR-423-5p, hsa-miR-451a, and hsa-miR-486-5p were differentially expressed in both LTB and active TB patients serum exosomes (Hashimoto et al., 2020). Lyu L et al., 2019 showed that four LTB-specific miRNAs, hsa-let-7e-5p, hsa-let-7d-5p, hsa-miR-450a-5p, and hsa-miR-140-5p and four active TB miRNAs, hsa-miR-1246, hsa-miR-2110, hsa-miR-370-3p, and hsa-miR-28-3p panel provide diagnostic markers for LTB and active TB in serum exosomes (Lyu et al., 2019). Though these studies use serum, PBMCs, and serum exosomes of the blood, they show discordance. Moreover, NGS limitations, such as sequencing bias, library preparation, and adapter dimer, may lead to discordance among studies, particularly low abundant miRNAs. Thus, metadata analysis helps to overcome the discordance findings among different studies and will provide potential biomarkers (Alimadadi et al., 2020). In this study, we aim to do a metadata analysis of small-RNA sequencing datasets of blood small-RNA sequencing data for identifying potential biomarkers in TB disease progression.

## Materials and Methods

### Dataset selection

To retrieve relevant TB-associated miRNA profiling data for the meta-analysis, a web-based search in the Sequence Read Archive (SRA) database (https://www.ncbi.nlm.nih.gov/sra) was performed using the keywords “tuberculosis,” “small-RNA sequencing,” and “microRNA.” In total, 13 small-RNA sequencing datasets were identified in the SRA database. The datasets were manually reviewed, and only those fulfilling the following criteria were included for further analysis, miRNA expression profiling by Next-Generation Sequencing from peripheral blood, serum, and serum exosome samples. Finally, a total of seven datasets, PRJNA625982, PRJNA625983, PRJNA542815, PRJNA629528, PRJNA489396, PRJNA510806, and PRJNA226734 (Table1), were selected for analysis. Extrapulmonary datasets, patients with other infections like HIV, cells infected with *MTB*, and ATT-treated LTB cases were excluded from this study (PRJNA735638, PRJNA270244, PRJNA248348, PRJNA489396, PRJNA608187, six ATT-treated LTB samples from PRJNA542815). One dataset (PRJEB36699) was excluded because of limited information on samples. The above seven datasets (131 samples) are separated into three sets: healthy controls, LTB, and active TB.

### Data processing and differential expression analysis

The workflow of our current data analysis is shown in Figure 1A. Raw data (FASTQ files) were downloaded from NCBI using the SRA Toolkit. The quality of raw reads was measured using FastQC version 0.11.9 (Brandine et al., 2019), adaptor reads and low-quality bases were eliminated by fastp version 0.23.0 (Chen et al., 2018), and reads with a length of < 16 and > 30 nt long were removed using BBMap/reformat.sh. Preprocessed reads were mapped with the human reference genome GRCh38 using STAR version 2.4.0.1 (Dobin et al., 2013) and then quantified using FeatureCounts version 2.0.3 (Liao et al., 2014). The read counts of less than 50 were removed from the data. miRNA read counts were normalized using TMM (Liu et al., 2019, Maza, 2016). All the samples selected for this study were combined by their miRNAID using an R-script. Principal component analysis (PCA) and clustering analysis were plotted using the R-script (Jafarzadegan et al., 2019). Differential expression (DE) analysis was performed by edgeR version 3.34.0 (Robinson et al., 2010). The miRNAs were considered significant expressions if the absolute fold change was >1.0, an adj. p <0.05, and log Counts per Million (CPM) greater than or equal to 2. Volcano plots were plotted using VolcaNoseR (Goedhart et al., 2020). The box plot of log CPM values with mean and Standard Error of Mean (SEM) values was calculated using the parametric t-test statistical analysis method in GraphPad Prism v8.0.2. A *P-value* of less than 0.05 was considered statistically significant. From that, 52 miRNAs were filtered based on TB-specific, active TB-specific, disease progressive, and LTB-specific miRNAs. The networks were constructed for both upregulated and downregulated miRNAs.

**Figure 1.**
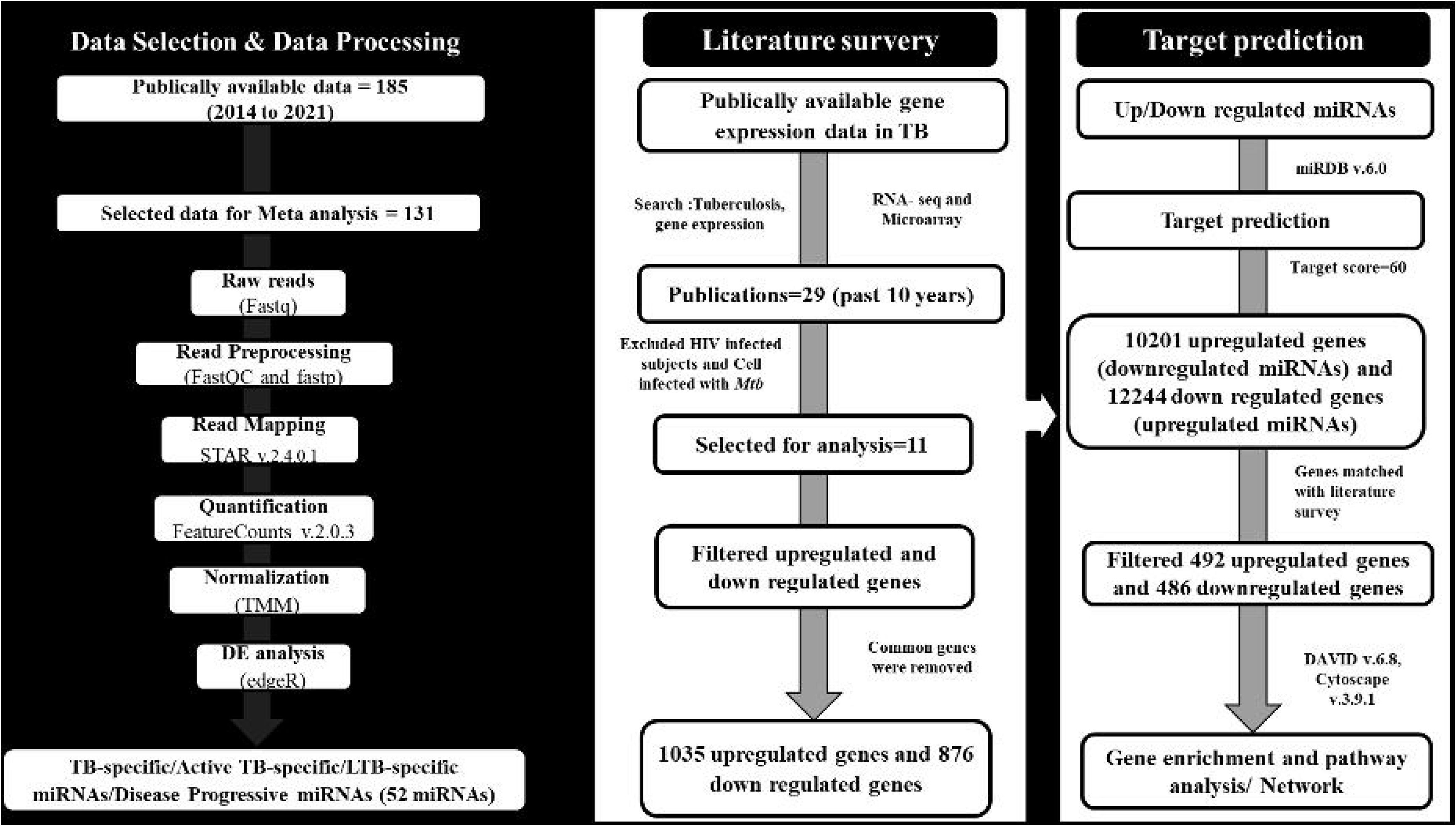
Schematic representation of workflow followed for metadata analysis. **(A)** miRNA data selection and processing of data **(B)** Transcriptome data selection, target prediction, and pathway analysis.

### miRNA Target prediction

Above 52 miRNAs were filtered as 21 upregulated and 31 downregulated miRNAs and target prediction was performed using miRDB v6.0 (Chen et al., 2020). In the miRDB database for target gene selection, we selected a prediction score of 60%. Duplicate genes were removed, and unique genes were filtered. The transcriptome gene expression data of tuberculosis pathogenesis were searched in PubMed using the following keywords “tuberculosis, gene expression” and filtered based on full text, books and documents, metadata analysis, review, systemic review, and the past 10 years. Eleven out of twenty-nine publications were identified and included in the gene selection. For patients with HIV infection, cell culture data were excluded. Common genes present in both upregulated and downregulated miRNAs were removed. Unique genes matched with predicted data of both upregulated and downregulated miRNAs were used for further analysis.

### miRNA-target functional network analysis

To predict the functions of the selected miRNAs in TB pathogenesis, we performed the functional enrichment analysis of target genes using the Database for Annotation, Visualization, and Integrated Discovery (DAVID 6.8) (Dennis et al., 2003). A *P*-value less than 0.05 and an FDR value < 0.9 were used as the threshold to select significant Gene ontology (GO) and Kyoto Encyclopedia of Genes and Genomes (KEGG) pathways. We used Cytoscape (version 3.9.1) (Otasek et al., 2019) software to construct the miRNA-target gene functional network with the TB-associated significantly enriched pathways of interest. miRNA and their target genes and pathway analysis workflow are shown in Figure 1B.

## Results

### Details of miRNA expression data

One hundred thirty-one samples were included in this study. From there, 45 controls (PRJNA625982 (13), PRJNA625983 (12), PRJNA489396 (3), PRJNA510806 (1), PRJNA226734 (2), and PRJNA542815 (14)), 47 active TB samples (PRJNA510806 (1), PRJNA625982 (8), PRJNA625983 (10), PRJNA629528 (16), PRJNA489396 (3), PRJNA226734 (1), and PRJNA542815 (8)), and 39 were LTB patient samples (PRJNA226734 (1), PRJNA629528 (16), PRJNA510806 (1), and PRJNA542815 (21)). miRNA expression data included in the metadata analysis were peripheral blood mononuclear cells (n = 92), serum (n = 4), and serum exosomes (n = 35) in patient samples (Table 1, Supplementary Table S1). The average number of reads, alignment rate, and percentage of mapped reads were shown in Table 2. After preprocessing and merging, data was normalized using the normalization methods. Before (Supplementary Figure S1A) and after (Supplementary Figure S1B) normalization, the whole data was plotted on the principal component analysis (PCA) plot. Further, it showed the apparent clustering of PBMC samples into two groups, and serum and serum exosomes were grouped into a single group (Supplementary Figure S2).

**Table 1.** Datasets for metadata analysis.

| Bioproject Accession ID | References | NO. of samples | Sample Type |
| --- | --- | --- | --- |
| PRJNA6259982 | Gu W et al., 2020 | 20 | PBMC |
| PRJNA6259983 | Gu W et al., 2020 | 23 | PBMC |
| PRJNA629528 | S Hashimoto et al., 2020 | 32 | Serum Exosomes |
| PRJNA542815 | de Araujo LS et al., 2019 | 43 | PBMC |
| PRJNA510806 | L Lyu et al., 2019 | 3 | Serum Exosomes |
| PRJNA489396 | F Liu et al., 2018 | 6 | PBMC |
| PRJNA226734 | Zhang H et al., 2014 | 4 | Serum |

Seven datasets (n = 131) were selected for metadata analysis.

**Table 2.** Data Statistics.

| Characteristics | Control | LTB | TB |
| --- | --- | --- | --- |
| NO. of samples | 45 | 39 | 47 |
| NO. of reads (in Millions) | 16.4 | 17.4 | 15.0 |
| % Of mapped reads | 99.06 | 96.5 | 95.9 |
| % Of Unmapped reads | 0.94 | 3.5 | 4.1 |
| Average mapped length | 24.9 | 22.7 | 23.2 |
| Avg NO. of miRNAs | 635 | 386 | 505 |

The average alignment rate of control, LTB, and active TB samples was 99.06%, 96.5%, and 95.9%, respectively.

### Differential expression of miRNAs in tuberculosis

Three hundred and thirty-three miRNAs were differentially expressed in active TB cases compared to healthy controls. From there, 222 miRNAs were upregulated, and 111 were downregulated miRNAs. The topmost differentially expressed miRNAs ordered by log2 fold change (FC) are presented in Table 3 and Figure 2. Four hundred and sixty-nine miRNAs were identified as differentially expressed in active TB compared with LTB samples (Supplementary Figure S3). Of these, 290 miRNAs were upregulated (those with a positive Log2 FC), and 179 were downregulated (those with a negative Log2 FC). Five hundred and thirteen miRNAs were identified as differentially expressed in LTB compared with healthy control samples (Supplementary Figure S4). Of these, 251 miRNAs were upregulated (those with a positive Log2 FC), and 262 were downregulated (those with a negative Log2 FC). We prioritized 52 miRNAs of which nine were TB-Specific (Supplementary Table S2, Supplementary Figure S5A) (Differentially expressed in both active TB and LTB cases compared to healthy controls and no significant difference between active and latent TB), eight active TB-specific miRNAs (Supplementary Table S3, Supplementary Figure S5B), seven were disease progressive miRNAs (Supplementary Table S4, Supplementary Figure S6), and 28 were LTB-specific miRNAs (Supplementary Table S5, Supplementary Figure S7). Among them, 31 miRNAs were downregulated miRNAs and 21 were upregulated miRNAs.

**Figure 2.**
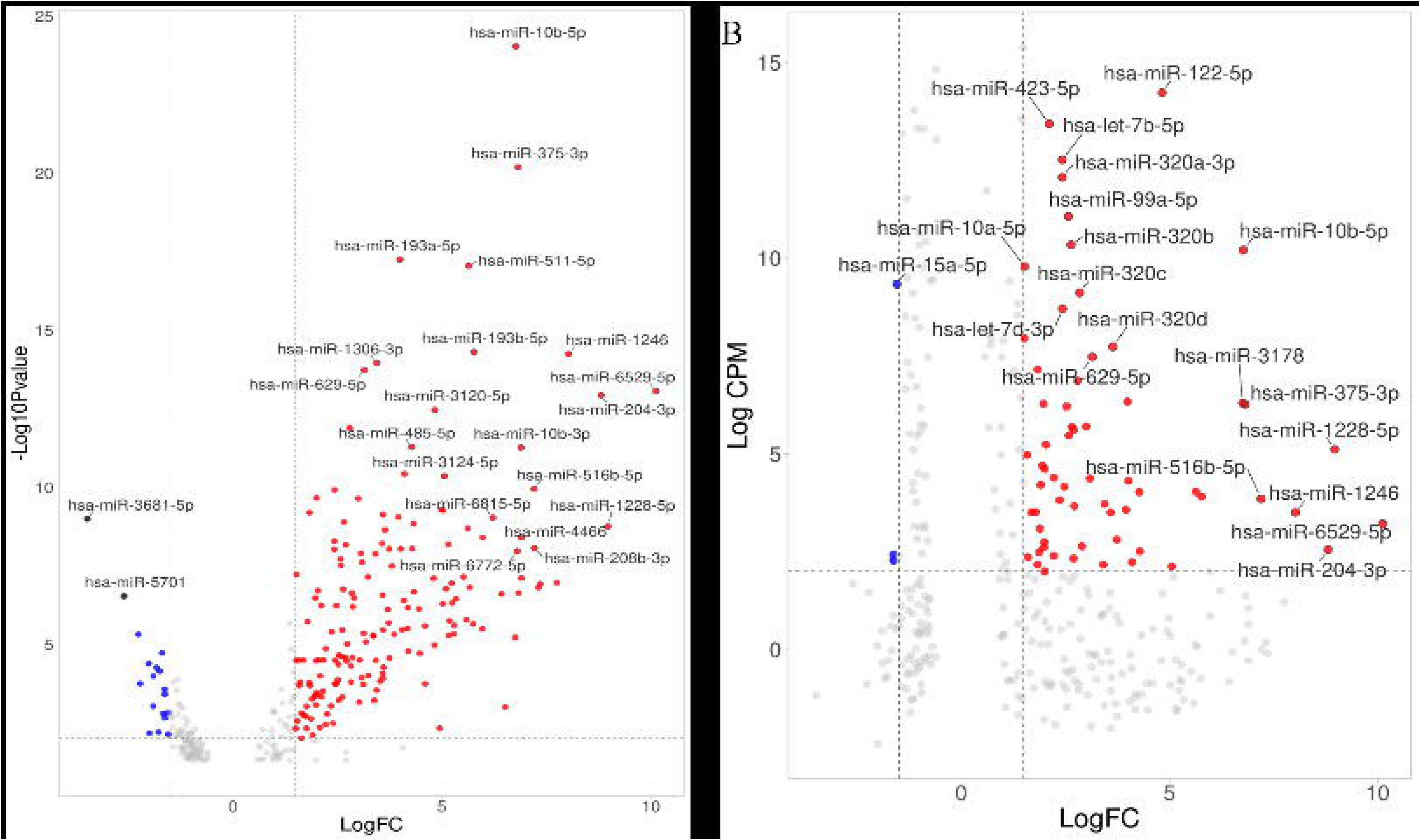
The volcano plot shows the differential expression of microRNAs in Active TB samples compared with healthy control samples. Red color represents upregulated miRNAs and blue color represents downregulated miRNAs. The top 20 miRNAs were labeled in the plot. We consider miRNAs significant based on absolute fold change was >1.0 and adj. p <0.05 and log CPM greater than or equal to 2. **(A)** The X-axis shows Log2 FC and the Y-axis shows the minus Log10 P-value. **(B)** The X-axis shows Log2 FC and the Y-axis shows the log CPM.

**Table 3.**
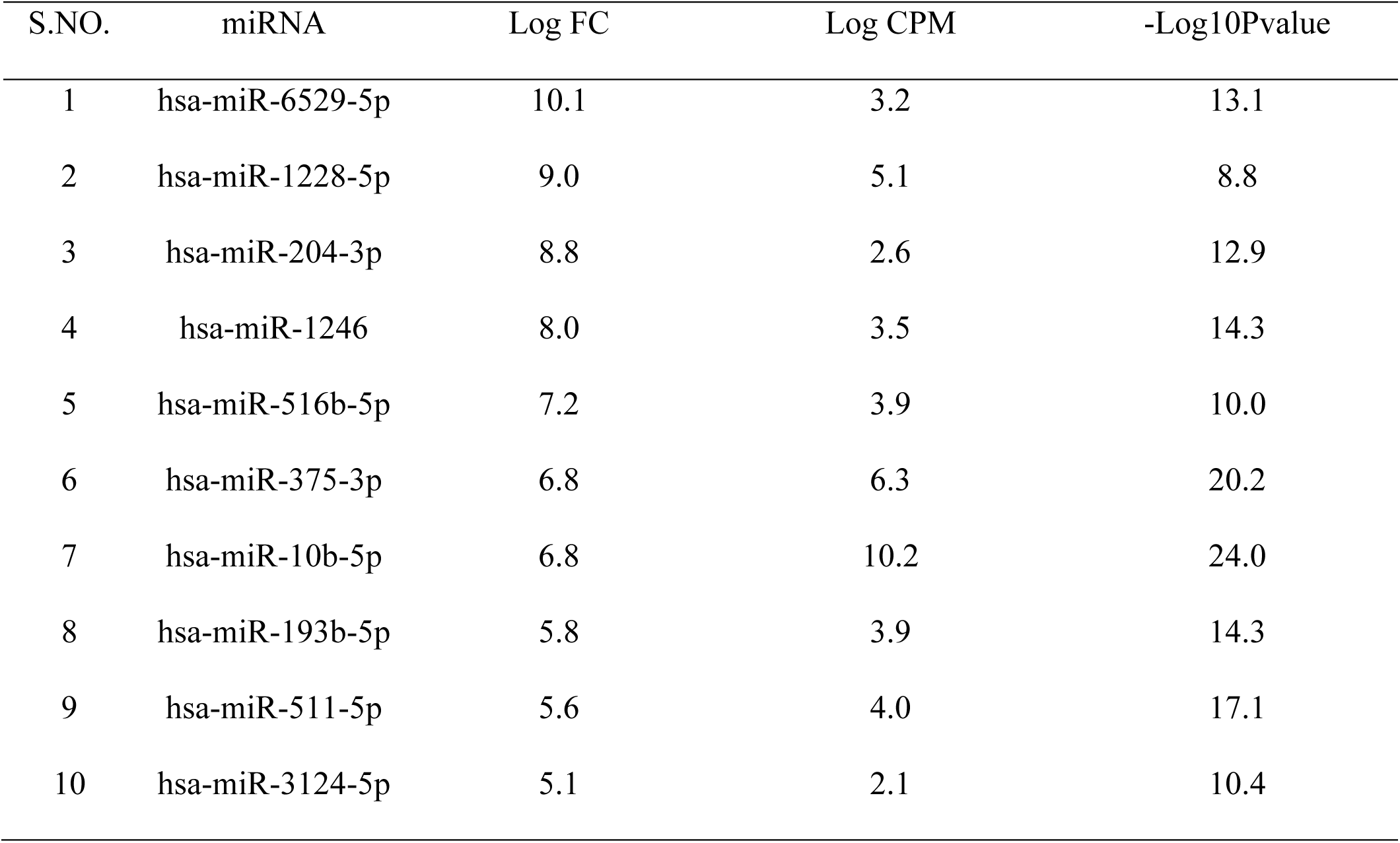
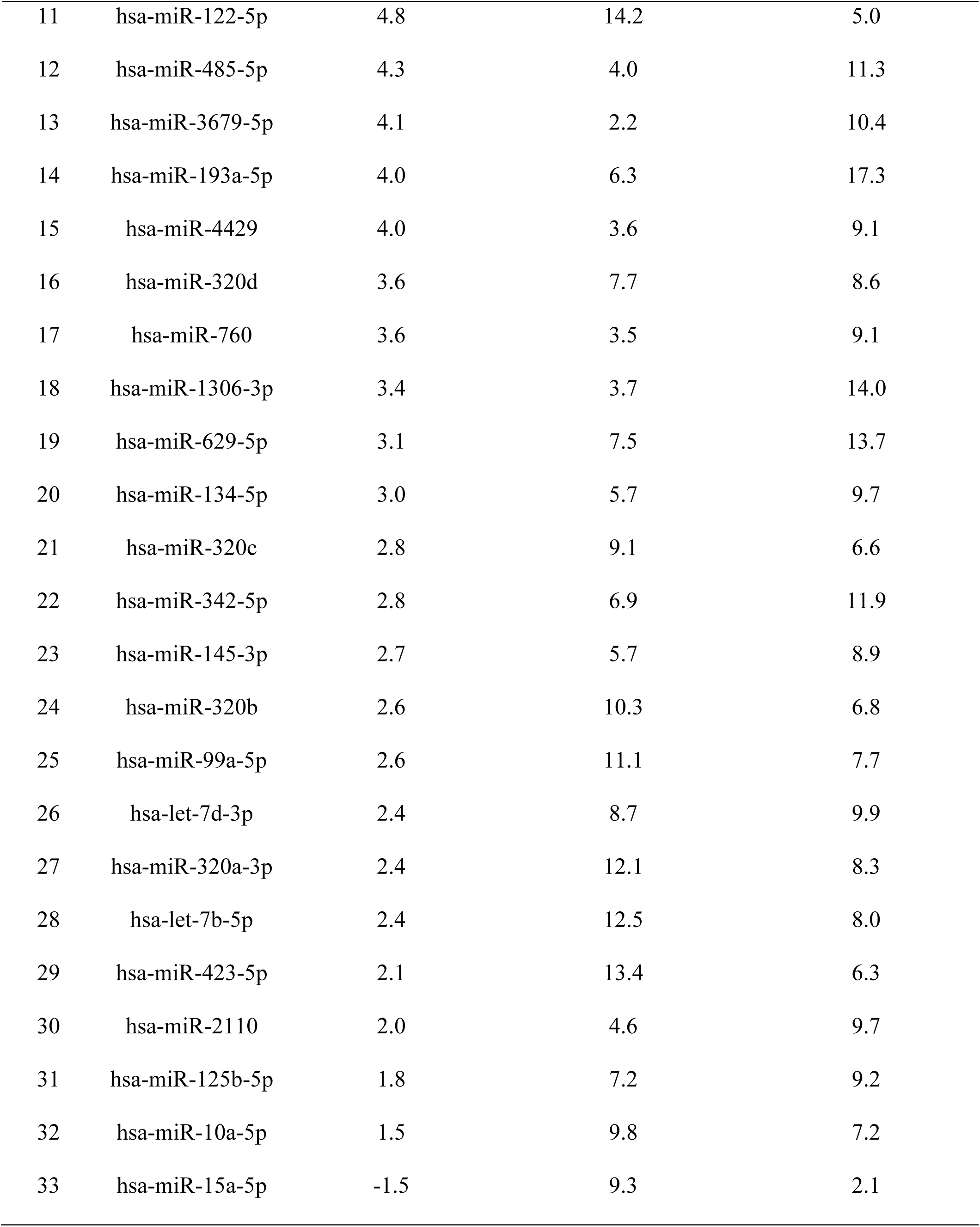
Differentially Expressed miRNAs in Active TB Compared to Controls.

| S.NO. | miRNA | Log FC | Log CPM | -Log10Pvalue |
| --- | --- | --- | --- | --- |
| 1 | hsa-miR-6529-5p | 10.1 | 3.2 | 13.1 |
| 2 | hsa-miR-1228-5p | 9.0 | 5.1 | 8.8 |
| 3 | hsa-miR-204-3p | 8.8 | 2.6 | 12.9 |
| 4 | hsa-miR-1246 | 8.0 | 3.5 | 14.3 |
| 5 | hsa-miR-516b-5p | 7.2 | 3.9 | 10.0 |
| 6 | hsa-miR-375-3p | 6.8 | 6.3 | 20.2 |
| 7 | hsa-miR-10b-5p | 6.8 | 10.2 | 24.0 |
| 8 | hsa-miR-193b-5p | 5.8 | 3.9 | 14.3 |
| 9 | hsa-miR-511-5p | 5.6 | 4.0 | 17.1 |
| 10 | hsa-miR-3124-5p | 5.1 | 2.1 | 10.4 |
| 11 | hsa-miR-122-5p | 4.8 | 14.2 | 5.0 |
| 12 | hsa-miR-485-5p | 4.3 | 4.0 | 11.3 |
| 13 | hsa-miR-3679-5p | 4.1 | 2.2 | 10.4 |
| 14 | hsa-miR-193a-5p | 4.0 | 6.3 | 17.3 |
| 15 | hsa-miR-4429 | 4.0 | 3.6 | 9.1 |
| 16 | hsa-miR-320d | 3.6 | 7.7 | 8.6 |
| 17 | hsa-miR-760 | 3.6 | 3.5 | 9.1 |
| 18 | hsa-miR-1306-3p | 3.4 | 3.7 | 14.0 |
| 19 | hsa-miR-629-5p | 3.1 | 7.5 | 13.7 |
| 20 | hsa-miR-134-5p | 3.0 | 5.7 | 9.7 |
| 21 | hsa-miR-320c | 2.8 | 9.1 | 6.6 |
| 22 | hsa-miR-342-5p | 2.8 | 6.9 | 11.9 |
| 23 | hsa-miR-145-3p | 2.7 | 5.7 | 8.9 |
| 24 | hsa-miR-320b | 2.6 | 10.3 | 6.8 |
| 25 | hsa-miR-99a-5p | 2.6 | 11.1 | 7.7 |
| 26 | hsa-let-7d-3p | 2.4 | 8.7 | 9.9 |
| 27 | hsa-miR-320a-3p | 2.4 | 12.1 | 8.3 |
| 28 | hsa-let-7b-5p | 2.4 | 12.5 | 8.0 |
| 29 | hsa-miR-423-5p | 2.1 | 13.4 | 6.3 |
| 30 | hsa-miR-2110 | 2.0 | 4.6 | 9.7 |
| 31 | hsa-miR-125b-5p | 1.8 | 7.2 | 9.2 |
| 32 | hsa-miR-10a-5p | 1.5 | 9.8 | 7.2 |
| 33 | hsa-miR-15a-5p | -1.5 | 9.3 | 2.1 |

### Functional analysis through miRNA-target-pathway network analysis

Thus, ten thousand two hundred and one target genes were predicted for 31 downregulated miRNAs. From the publicly available transcriptome gene expression data (Supplementary Table S10, S11, and S12), thousand and thirty-five upregulated genes were identified. Four hundred and ninety-two genes were matched in both groups. Eight hundred seventy-six genes were predicted for 21 upregulated miRNAs and 486 genes were matched with gene expression data (Supplementary Figure 2B). Pathway enrichment analysis was done for both up and downregulated miRNAs. Common pathways were excluded for further analysis. Downregulated miRNA’s target genes showed an association of 233 GOTERM_BP_DIRECT pathways and 268 KEGG signaling pathways. Supplementary Table S6 and S7 show the top pathways associated with GO and KEGG pathways, respectively. Downregulated miRNAs GO categories were reported as inflammatory response, cellular response to interferon-gamma, innate immune response, response to hypoxia, cellular response to lipopolysaccharides, extracellular matrix organization, leukocyte cell-cell adhesion, and positive regulation of nitric oxide biosynthetic process, etc., (Figure 3A). TNF signaling pathway, phagosome, PI3K-Akt signaling pathway, NOD-like receptor signaling pathway, etc., were downregulated miRNAs KEGG pathways of our interest (Figure 3B). Four hundred and eighty-six downregulated genes were matched with transcriptome data of 21 upregulated miRNAs. Pathway enrichment analysis using downregulated target genes showed an association of 125 GOTERM_BP_DIRECT pathways and 17 KEGG signaling pathways (Supplementary Tables S8 and S9). All upregulated miRNAs target genes GO categories have a role in protein autophosphorylation, positive regulation of angiogenesis, B-cell activation, negative regulation of cell proliferation, negative thymic T-cell selection, response to growth factor, T-cell activation, integrin-mediated signaling pathway, intracellular signal transduction, CD8-positive, alpha-beta T cell lineage commitment, response to bacterium, and angiogenesis, etc., (Figure 4A). NF-kappa B signaling pathway, and allograft rejection, KEGG pathways of our interest were significantly downregulated in TB pathogenesis (Figure 4B). From the above miRNAs, we further filtered eight miRNAs (six downregulated miRNAs and two upregulated miRNAs) based on their high fold change (log FC≥4) and shared target genes and pathways (Supplementary Figure S8 and Supplementary Figure S9).

**Figure 3.**
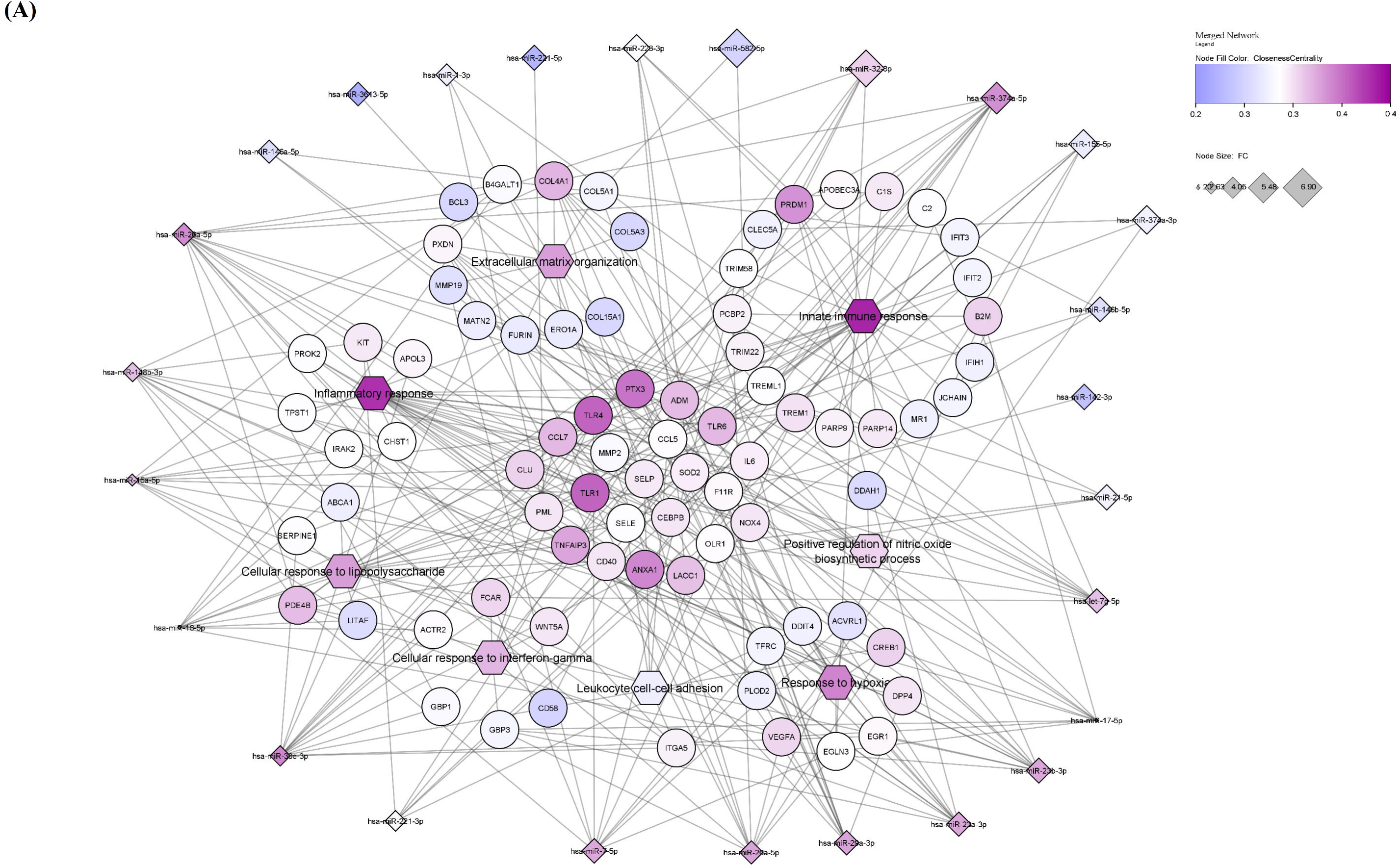
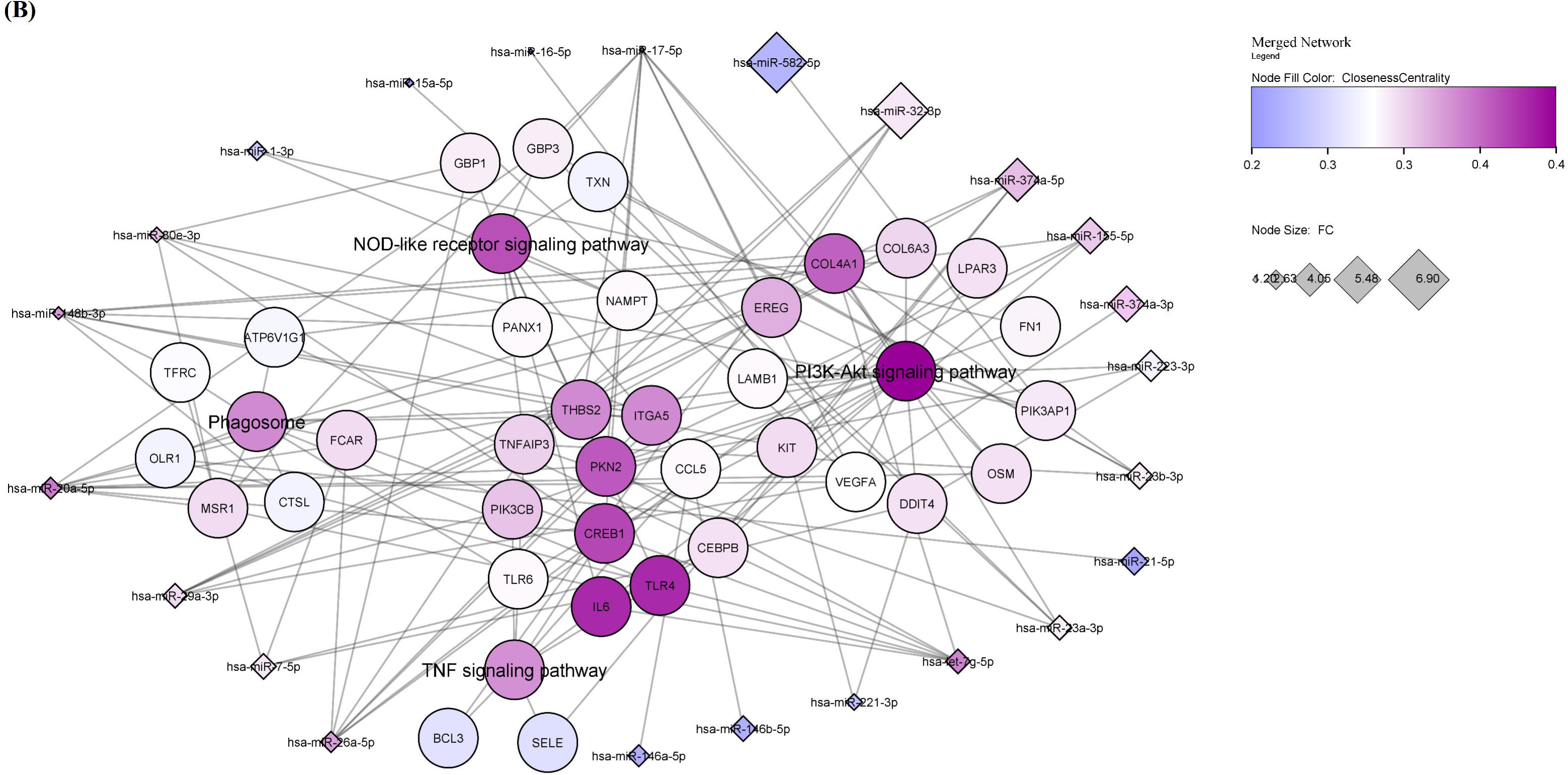
Network of upregulated pathways. Downregulated miRNAs and their target genes having higher closeness centrality were marked with a blue to purple color circle; the intensity of the color corresponds to the level of closeness centrality. Diamond shape-downregulated miRNAs (low fold change to high fold change were marked as blue to purple color), hexagonal shape-pathways, and circle shape-target genes. **(A)** GO Biological pathways **(B)** KEGG pathways.

**Figure 4.**
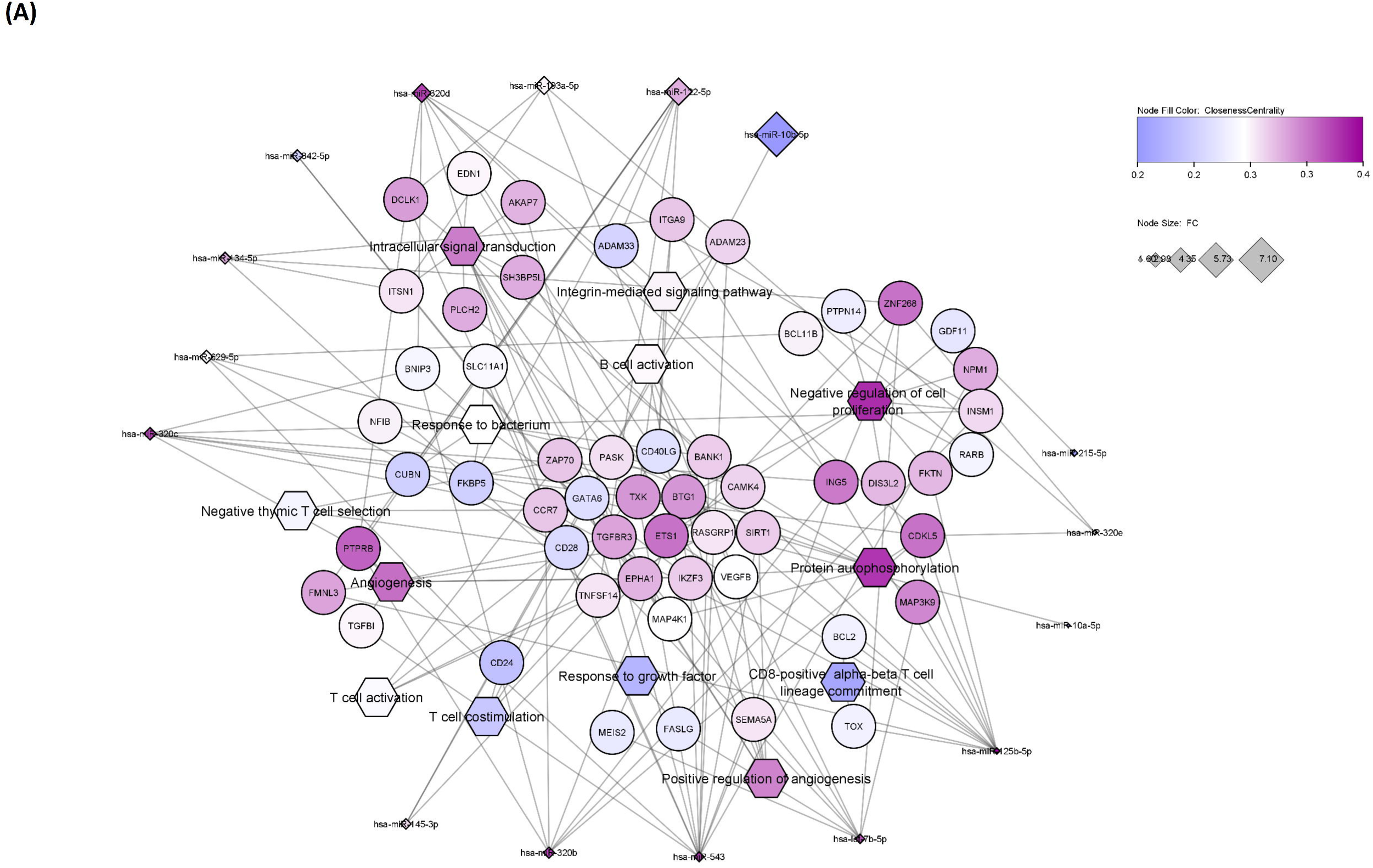
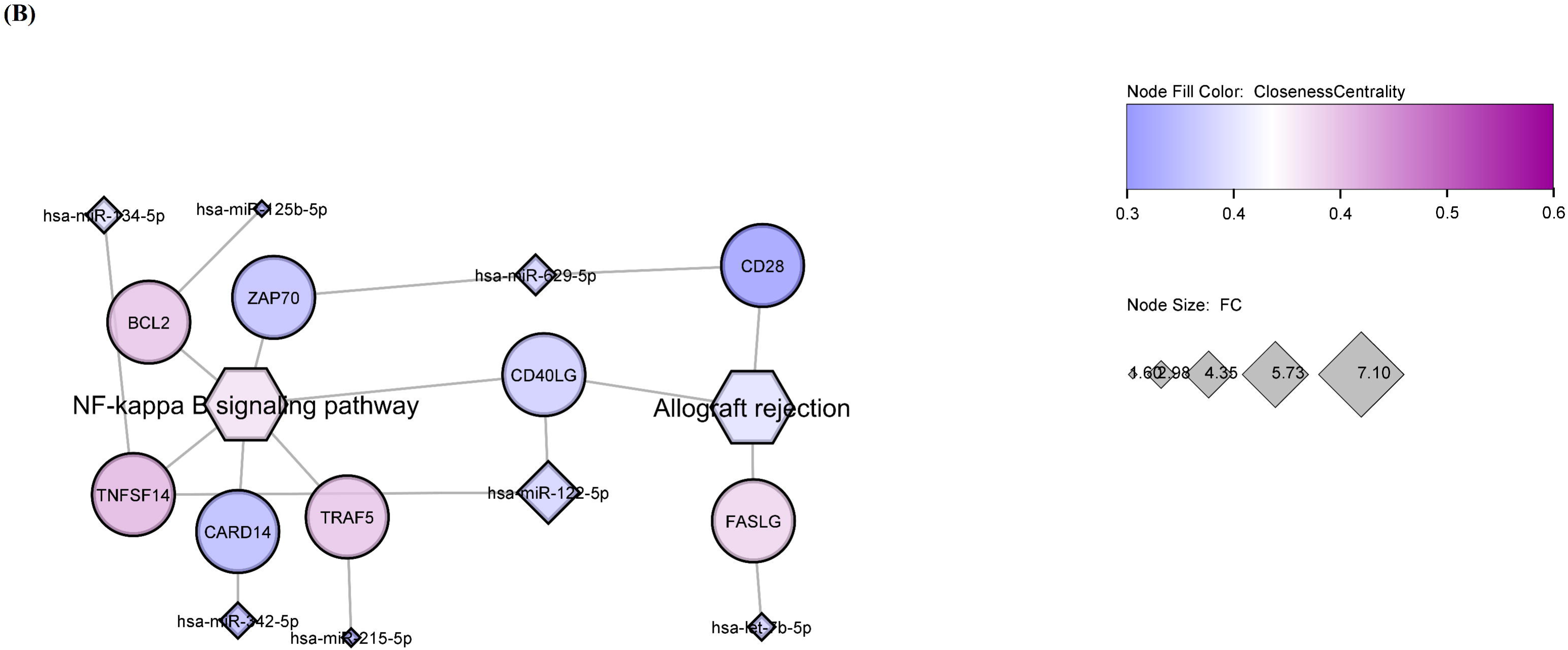
Network of Downregulated pathways. Upregulated miRNAs and their target genes having higher closeness centrality were marked with a blue to purple color circle; the intensity of the color corresponds to the level of closeness centrality. Diamond shape-downregulated miRNAs (low fold change to high fold change were marked as blue to purple color), hexagonal shape--pathways, and circle shape-target genes. **(A)** GO Biological pathways **(B)** KEGG pathways.

## Discussion

Metadata analysis always helps to increase the statistical power and provide more robust and reliable conclusions than individual studies. Hence, we aim to identify miRNAs in TB, to do that, we have found miRNA expression from PBMC, serum, and serum exosome data in tuberculosis. From that, we prioritized 52 miRNAs of which nine were TB-specific, eight were active TB-specific miRNAs, seven were disease-progressive miRNAs, and 28 were LTB-specific miRNAs. Among them, 31 miRNAs were downregulated and 21 miRNAs were upregulated.

Hashimoto et al.,2020 (PRJNA629528) showed that hsa-miR-21-5p was upregulated in active TB compared to LTB samples, but in our metadata analysis, hsa-miR-21-5p was downregulated, and hsa-miR-122-5p was upregulated in both LTB and active TB compared to healthy controls. Lyu et al., 2019 (PRJNA510806) identified miRNAs, hsa-miR-450a-5p, and hsa-miR-140-5p in LTB were downregulated in our data. In contrast to Lyu et al., 2019, hsa-miR-29a-3p, and hsa-miR-146b-5p were downregulated in our data analysis. hsa-miR-629-5p was upregulated and hsa-miR-26a-5p was downregulated in both active TB and LTB samples in our analysis (Lyu et al., 2019).

Active TB-specific miRNA, hsa-miR-320d is an anti-inflammatory miRNA upregulated in our data (Faiz et al., 2015). Other miRNAs, hsa-miR-193a-5p, hsa-miR-10a-5p, and hsa-miR-10b-5p, significantly target various cytokines such as TNF-α, IFNγ, IL-1β, IL-8, and IL-10 have a role in TB pathogenesis (Ulger et al., 2022). Both TB-specific and active TB miRNAs served as TB-susceptible (Increases severity of TB disease) miRNAs. TB-specific upregulated hsa-miR-122-5p target gene, CD40 ligand alone, and combination with IFN**γ** activates the vitamin D-dependent antimicrobial pathway and triggers antimicrobial activity against intracellular *MTB* infection in T cells have a role in NF-kappa signaling pathway and T cell co-stimulation (Klug-Micu et al., 2013). In tuberculosis infection, *MTB* cell wall glycolipids, especially mannose-capped lipoarabinomannan (ManLAM), inhibit T cell receptor signaling through the suppression of ZAP-70, which is a target gene of miR-629-5p (Mahon et al., 2012). The CD28, another target gene of hsa-miR-629-5p, was downregulated in progressors (TB infection to TB disease) compared to controls (Scriba et al., 2017). Genome-Wide Association Studies showed that hsa-miR-10a-5p and hsa-miR-10b-5p target gene GATA6 was a TB susceptible gene due to proximity to a susceptibility locus for TB on chromosome 18q11.2 variant (SNP rs4331426) (Zheng et al., 2018). In mouse experiments, C57BL/6 CCR7^−/−^ mice showed more susceptibility for TB compared to CCR7^+/+^ because of a deficiency of CCR7, which is a target for hsa-miR-320b. Active TB-specific miRNA, hsa-miR-let-7b-5p was upregulated in H37Rv-infected THP-1 macrophages, promoting mycobacterial survival through the degradation of *Fas* transcripts and inhibiting apoptosis (Sampath et al., 2021, Tripathi et al., 2018). In our study, hsa-miR-320a-3p expression was upregulated in active TB cases compared to healthy controls. The validated high expression levels of hsa-miR-320a were reported in pan-susceptible TB patients than in drug-resistant TB patients (Cui et al., 2017). Cui et al. reported that hsa-miR-320a plays a role in the modulation of different cytokine production in TB. Decreased expression of hsa-miR-320a may play a role in reactivating cell migration and proliferation in the lung tissue in TB disease progression in drug-resistant cases (Cui et al., 2017). hsa-miR-125b-5p target gene BCL2 polymorphisms may be associated with susceptibility to both active and LTB (He et al., 2022). hsa-miR-543 target gene TXK autophosphorylation, active form promotes IFNγ synthesis (Kashiwakura et al., 2002). SIRT1 activator inhibited *MTB*-induced apoptosis in mouse peritoneal macrophages, which was a target gene of hsa-miR-543 (Yang et al., 2020). Upregulated hsa-miR-193a-5p target gene VEGF has a role in triggering angiogenesis and vascularization of lesions (Maison, 2022). hsa-miR-134-5p target gene *ITSN1* differentiates TB disease from infection peripartum, which has a role in adaptive inflammatory responses, including CD4^+^ T-cell activation (Mathad et al., 2022).

LTB-specific hsa-miR-582-5p target gene *COL5A1* polymorphisms implicated as genetic markers for pulmonary function (Zhetkenev et al., 2021). hsa-miR-374a-5p target *CREBPB* gene phosphorylates CREB and promotes IFN**γ** in T lymphocytes while favoring M2 polarization during *MTB* infection (Wen et al., 2021). Monocytes CD14+ CD16+sub type expression was higher in TST-positive individuals than active TB patients or healthy controls with negative TST, showing that these cells contribute innate immunity against TB infection. monocytes surface marker CLEC5A gene was targeted by hsa-miR-374a-3p; thus, the monocyte and lymphocyte ratio suggests that TST may risk for developing TB in contacts (Sampath et al., 2018). hsa-miR-1-3p target gene *MR1* in the control of *MTB* suggested that an MR1 single-nucleotide polymorphism was associated with Mtb meningitis (Kim et al., 2022). TLR-4 mutational study showed that hsa-miR-7-5p target gene TLR4 mutation is associated with increased susceptibility to TB disease (Thada et al., 2021).

miR-17 family members such as hsa-miR-17-5p (TB-specific) and hsa-miR-20a-5p (LTB-specific) target gene CREB1 proteins levels were increased in LTB patients compared to active TB leads to the increased IFNγ production to control the *MTB* growth (Liu et al., 2010). hsa-miR-16-5p and hsa-miR-15a-5p target gene TLR1 polymorphisms may be associated with a high risk of developing TB disease (Zhou et al., 2019). hsa-miR-32-3p, hsa-miR-374a-5p (Wang et al., 2022), hsa-miR-223-3p (Sinigaglia et al., 2020), hsa-miR-155-5p (Liu et al., 2023), and hsa-miR-374a-3p (Wang et al., 2022) were control the TB through TNF signaling pathways. miR-122-5p distinguishes LTB from active TB with an accuracy of 71.8% having a role in tuberculosis through different pathways (Supplementary Figure S9) (Godkowicz et al., 2022).

We identified several TB-specific miRNAs, LTB-specific, active TB, and disease-progressive miRNAs. These miRNAs may play an important role in TB disease progression from LTB to TB. Further, eight miRNAs, hsa-miR-155-5p, hsa-miR-223-3p, hsa-miR-32-3p, hsa-miR-374a-3p, hsa-miR-374a-5p, hsa-miR-582-5p, hsa-miR-320d, and miR-122-5p were selected. These, eight miRNAs could serve as biomarkers based on their role in TB pathogenesis through PI3K-Akt signaling pathway, TNF-signaling pathway, phagosome, and NOD-like signaling pathway, which required further validation.

## Data Availability

All data produced in the present work are contained in the manuscript

## Authors’ contribution

**Swathi Chadalawada**: Data curation, Data analysis, Writing-Reviewing, and Editing, **Bharanidharan Devarajan:** Conceptualization, Reviewing, and Editing, and **SR. Rathinam:** Reviewing and Editing.

## Acknowledgments

None

## Funding Information

Senior Research Fellow -Indian Council of Medical Research (ICMR), India (Sanction letter No. 2019-4013/Gen-BMS dt.30.09.2019).

## Ethics declarations

### Conflict of interest

**SC**, None; **BD**, None; **SRR**, None.

### Ethical Approval

Not Applicable.

### Availability of data and materials

PRJNA6259982, PRJNA6259983, PRJNA629528, PRJNA542815, PRJNA510806, PRJNA489396, and PRJNA226734.

### Consent to participate

Not applicable.

### Consent to publish

Not applicable.

## Supplementary Information

**Supplementary Figure S1. PCA plot for meta-analysis. (A)** Before Normalization. **(B)** After Normalization

**Supplementary Figure S2. Clustering tree After Normalization.** After Normalization, clear clustering of PBMC samples was into two groups, and serum and Serum exosomes were grouped into a single. P-PBMC; S – Serum; SE – Serum Exosomes

**Supplementary Figure S3. The volcano plot shows the differential expression of microRNAs in Active TB samples compared with LTB.** Red color represents upregulated miRNAs and blue color represents downregulated miRNAs. The top 20 miRNAs were labeled in the plot. We consider miRNAs significant based on absolute fold change was >1.0 and adj. p <0.05 and log CPM greater than or equal to 2. **(A)** The X-axis shows the Log2 fold change, and the Y-axis shows the minus Log10 P-value. **(B)** The X-axis shows the Log2 fold change, and the Y-axis shows the log CPM. We consider miRNAs significant based on absolute fold change was >1.0 and adj. p < 0.05 and log CPM greater than or equal to 2.

**Supplementary Figure S4. The volcano plot shows the differential expression of microRNAs in LTB samples compared with healthy control samples**. Red color represents upregulated miRNAs and blue color represents downregulated miRNAs. The top 20 miRNAs were labeled in the plot. We consider miRNAs significant based on absolute fold change was >1.0 and adj. p <0.05 and log CPM greater than or equal to 2. **(A)** The X-axis shows the Log2 fold change, and the Y-axis shows the minus Log10 P-value. **(B)** The X-axis shows the Log2 fold change, and the Y-axis shows the log CPM.

**Supplementary Figure S5. The bar graphs were plotted for miRNAs in TB-specific and active TB.** CPM - Counts per million. A parametric t-test statistical analysis method was used for data significance. *P<0.05, ** P<0.01, and ***P<0.001. **(A)** TB-specific miRNAs **(B)** Active TB miRNAs

**Supplementary Figure S6. The bar graphs were plotted for miRNAs in TB disease progression.** The progressive differential miRNA expression was identified in TB compared to healthy controls. CPM - Counts per million. A parametric t-test statistical analysis method was used for data significance. *P<0.05, ** P<0.01, and ***P<0.001.

**Supplementary Figure S7. Bar graphs were plotted for both LTB-specific miRNAs in blood.** The differential expression of miRNAs was identified in LTB compared to healthy controls and active TB. CPM - Counts per million. A parametric t-test statistical analysis method was used for data significance. *P<0.05, ** P<0.01, ***P<0.001, and ****P<0.0001.

**Supplementary Figure S8. Network of six downregulated miRNAs.** Downregulated miRNAs and their shared common target genes having higher closeness centrality were marked with a blue to purple color circle; the intensity of the color corresponds to the level of closeness centrality. Diamond shape-downregulated miRNAs. hexagonal shape-pathways, and circle shape-target genes.

**Supplementary Figure S9. Network of upregulated miRNAs.** Two upregulated miRNAs and their shared common target genes having higher closeness centrality were marked with a blue to purple color circle; the intensity of the color corresponds to the level of closeness centrality. Diamond shape-downregulated miRNAs (low fold change to high fold change were marked as blue to purple color), hexagonal shape-pathways, and circle shape-target genes

## Notes

### Competing Interest Statement

The authors have declared no competing interest.

### Funding Statement

This study did not receive any funding

## References

L. Ai et al. Clinical value of interferon-γ release assay in the diagnosis of active tuberculosis. Exp. Ther. Med. (2019) 18(2):1253–1257. 10.3892/etm.2019.7696.

A. Alimadadi et al. Identification of Upstream Transcriptional Regulators of Ischemic Cardiomyopathy Using Cardiac RNA-Seq Meta-Analysis. Int. J. Mol. Sci. (2020) 21(10):3472. 10.3390/ijms21103472.

A. Behrouzi et al. The role of host miRNAs on Mycobacterium tuberculosis. ExRNA. (2019) 1. 10.1186/s41544-019-0040-y.

G. Brandine et al. Falco: high-speed FastQC emulation for quality control of sequencing data. F1000Res. (2019) 8:1874. 10.12688/f1000research.21142.2.

S. Chakrabarty et al. Host and MTB genome encoded miRNA markers for diagnosis of tuberculosis. Tuberculosis (Edinb). (2019) 116:37–43. 10.1016/j.tube.2019.04.002.

S. Chen et al. fastp: an ultra-fast all-in-one FASTQ preprocessor. Bioinformatics (2018) 34(17):i884–i890. 10.1093/bioinformatics/bty560.

Y. Chen et al. miRDB: an online database for the prediction of functional microRNA targets. Nucleic Acids Res. (2020) 48:D127–D31. 10.1093/nar/gkz757.

S.B. Cohen et al. Alveolar Macrophages Provide an Early Mycobacterium tuberculosis Niche and Initiate Dissemination. Cell Host Microbe. (2018) 24:439–46.e4. 10.1016/j.chom.2018.08.001.

J.Y. Cui et al. Characterization of a novel panel of plasma microRNAs that discriminates between Mycobacterium tuberculosis infection and healthy individuals. PloS One (2017) 12(9):e0184113. 10.1371/journal.pone.0184113.

L.S. de Araujo et al. Reprogramming of Small Noncoding RNA Populations in Peripheral Blood Reveals Host Biomarkers for Latent and Active Mycobacterium tuberculosis Infection. MBio (2019) 10(6):e01037–19. 10.1128/mBio.01037-19.

L.S. de Araujo et al. Transcriptomic Biomarkers for Tuberculosis: Validation of NPC2 as a Single mRNA Biomarker to Diagnose TB, Predict Disease Progression, and Monitor Treatment Response. Cells (2021) 10(10):2704. 10.3390/cells10102704.

N.B.R. de Sá et al. Clinical and genetic markers associated with tuberculosis, HIV-1 infection, and TB/HIV-immune reconstitution inflammatory syndrome outcomes. BMC Infect. Dis. (2020) 20(1):59. 10.1186/s12879-020-4786-5.

G. Dennis et al. DAVID: Database for Annotation, Visualization, and Integrated Discovery. Genome Biol. (2003) 4:P3.

A. Dobin et al STAR: ultrafast universal RNA-seq aligner. Bioinformatics (2013) 29(1):15–21. 10.1093/bioinformatics/bts635.

R. Domingo-Gonzalez et al. Cytokines and Chemokines in Mycobacterium tuberculosis Infection. Microbiol. Spectr. (2016) 4(5). 10.1128/microbiolspec.TBTB2-0018-2016.

A. Faiz et al. MiR-320d: A novel anti-inflammatory miRNA up regulated by corticosteroids. Eur. Respir. J. (2015) 46:OA2927. 10.1183/13993003.congress-2015.OA2927

M. Godkowicz et al. NOD1, NOD2, and NLRC5 Receptors in Antiviral and Antimycobacterial Immunity. Vaccines (Basel). (2022) 10(9):1487. 10.3390/vaccines10091487.

J.A-O. Goedhart et al. VolcaNoseR is a web app for creating, exploring, labeling and sharing volcano plots. Sci. Rep. (2020) 10(1):20560. 10.1038/s41598-020-76603-3.

S. Hashimoto et al. Developing a diagnostic method for latent tuberculosis infection using circulating miRNA. Transl. Med. Commun. (2020) 5,25. 10.1186/s41231-020-00078-7.

J.A-O. He et al Polymorphisms of the BCL2 gene associated with susceptibility to tuberculosis. Rev. Inst. Med. Trop. Sao Paulo (2022) 64:e59. 10.1590/S1678-9946202264059.

M. Jafarzadegan et al. Combining hierarchical clustering approaches using the PCA method. Expert. Syst. Appl. (2019) 137:1–10. 10.1016/j.eswa.2019.06.064.

J. Kashiwakura et al. Evidence of autophosphorylation in Txk: Y91 is an autophosphorylation site. Biol. Pharm. Bull. (2002) 25(6):718–21. 10.1248/bpb.25.718.

S-J. Kim et al. MR1- and HLA-E-Dependent Antigen Presentation of Mycobacterium tuberculosis. Int. J. Mol. Sci. (2022) 23(22):14412. 10.3390/ijms232214412.

G.M. Klug-Micu et al. CD40 ligand and interferon-γ induce an antimicrobial response against Mycobacterium tuberculosis in human monocytes. Immunology (2013) 139(1):121–8. 10.1111/imm.12062.

S.W. Lee et al. Gene expression profiling identifies candidate biomarkers for active and latent tuberculosis. BMC Bioinform. (2016) 17 Suppl 1(Suppl 1):3. 10.1186/s12859-015-0848-x.

Y. Liao et al. featureCounts: an efficient general purpose program for assigning sequence reads to genomic features. Bioinformatics (2014) 30(7):923–930. 10.1093/bioinformatics/btt656.

F. Liu et al. MicroRNA-27a controls the intracellular survival of Mycobacterium tuberculosis by regulating calcium-associated autophagy. Nat. Commun. (2018) 9(1):4295. 10.1038/s41467-018-06836-4.

J. Liu et al. Novel Biomarker Panel of Let-7d-5p and MiR-140-5p Can Distinguish Latent Tuberculosis Infection from Active Tuberculosis Patients. Infect. Drug. Resist. (2023) 16:3847–3859. 10.2147/IDR.S412116.

X. Liu et al. Normalization Methods for the Analysis of Unbalanced Transcriptome Data: A Review. Front. Bioeng. Biotechnol. (2019) 7:358. 10.3389/fbioe.2019.00358.

Y. Liu et al. CREB Is a Positive Transcriptional Regulator of Gamma Interferon in Latent but Not Active Tuberculosis Infections. Clin. Vaccine Immunol. (2010) 17(9):1377–80. 10.1128/CVI.00242-10.

L. Lyu et al. Small RNA Profiles of Serum Exosomes Derived From Individuals With Latent and Active Tuberculosis. Front Microbiol. (2019) 10:1174. 10.3389/fmicb.2019.01174.

R.N. Mahon et al. Mycobacterium tuberculosis ManLAM inhibits T-cell-receptor signaling by interference with ZAP-70, Lck and LAT phosphorylation. Cell Immunol. (2012) 275(1-2):98–105. 10.1016/j.cellimm.2012.02.009.

D.P. Maison. Tuberculosis pathophysiology and anti-VEGF intervention. J. Clin. Tuberc. Other Mycobact. Dis. (2022) 27: 100300. 10.1016/j.jctube.2022.100300.

J.S. Mathad et al. Transcriptional Analysis for Tuberculosis in Pregnant Women From the PRegnancy Associated Changes In Tuberculosis Immunology (PRACHITi) Study. Clin. Infect. Dis. (2022) 75(12):2239–2242. 10.1093/cid/ciac437.

E. Maza. In Papyro Comparison of TMM (edgeR), RLE (DESeq2), and MRN Normalization Methods for a Simple Two-Conditions-Without-Replicates RNA-Seq Experimental Design. Front Genet. (2016) 7:164. 10.3389/fgene.2016.00164.

M.D.B. Ndiaye et al. Plasma host protein signatures correlating with Mycobacterium tuberculosis activity prior to and during antituberculosis treatment. Sci. Rep. (2022) 12(1):20640. 10.1038/s41598-022-25236-9.

D. Otasek et al. Cytoscape Automation: empowering workflow-based network analysis. Genome Biol. (2019) 20(1):185. 10.1186/s13059-019-1758-4.

M. Ratti et al. MicroRNAs (miRNAs) and Long Non-Coding RNAs (lncRNAs) as New Tools for Cancer Therapy: First Steps from Bench to Bedside. Target Oncol. (2020) 15(3):261–278. 10.1007/s11523-020-00717-x.

M.D. Robinson et al. edgeR: a Bioconductor package for differential expression analysis of digital gene expression data. Bioinformatics (2010) 26(1):139–140. 10.1093/bioinformatics/btp616.

N. Sabir et al. miRNAs in Tuberculosis: New Avenues for Diagnosis and Host-Directed Therapy. Front Microbiol. (2018) 9:602. 10.3389/fmicb.2018.00602.

P. Sampath et al. Monocyte Subsets: Phenotypes and Function in Tuberculosis Infection. Front Immunol. (2018) 9:1726. 10.3389/fimmu.2018.01726.

P. Sampath et al. Monocyte and Macrophage miRNA: Potent Biomarker and Target for Host-Directed Therapy for Tuberculosis. Front Immunol. (2021) 12:667206. 10.3389/fimmu.2021.667206.

T.A-O. Scriba et al. Sequential inflammatory processes define human progression from M. tuberculosis infection to tuberculosis disease. PloS Pathog. (2017) 13(11):e1006687. 10.1371/journal.ppat.1006687.

A. Sinigaglia et al. Tuberculosis-Associated MicroRNAs: From Pathogenesis to Disease Biomarkers. Cells (2020) 9(10):2160. 10.3390/cells9102160.

S. Thada et al. Interaction of TLR4 and TLR8 in the Innate Immune Response against Mycobacterium Tuberculosis. Int. J. Mol. Sci. (2021) 22(4):1560. 10.3390/ijms22041560.

A. Tripathi et al. hsa-let-7b-5p facilitates Mycobacterium tuberculosis survival in THP-1 human macrophages by Fas downregulation. FEMS Microbiol. Lett. (2018) 365(7):fny040. 10.1093/femsle/fny040.

M. Ulger et al. Possible relation between expression of circulating microRNA and plasma cytokine levels in cases of pulmonary tuberculosis. J. Infect. Dev. Ctries. (2022) 16(7):1166–1173. 10.3855/jidc.15831.

L. Wang et al. MicroRNAs as immune regulators and biomarkers in tuberculosis. Front Immunol. (2022) 13:1027472. 10.3389/fimmu.2022.1027472.

Z. Wei, et al. The meta-analysis for ideal cytokines to distinguish the latent and active TB infection. BMC Pulm. Med. (2020) 20(1):248. 10.1186/s12890-020-01280-x.

Q. Wen et al. β-Arrestin 2 Regulates Inflammatory Responses against Mycobacterium tuberculosis Infection through ERK1/2 Signaling. J. Immunol. (2021) 206(11):2623–2637. 10.4049/jimmunol.2001346.

H. Yang et al. Sirtuin inhibits M. tuberculosis-induced apoptosis in macrophage through glycogen synthase kinase-3β. Arch. Biochem. Biophys. (2020) 694:108612.

Y. Yu et al. Different patterns of cytokines and chemokines combined with IFN-gamma production reflect Mycobacterium tuberculosis infection and disease. PLoS One (2012) 7:e44944. 10.1371/journal.pone.0044944

H. Zhang et al. Identification of serum microRNA biomarkers for tuberculosis using RNA-seq. PloS one (2014) 9:e88909. 10.1371/journal.pone.0088909.

R. Zheng et al. Genome-wide association study identifies two risk loci for tuberculosis in Han Chinese. Nat. Commun. (2018) 9(1):4072. 10.1038/s41467-018-06539-w.

S. Zhetkenev et al. Association of rs12722 COL5A1 with pulmonary tuberculosis: a preliminary case-control study in a Kazakhstani population. Mol. Bio. Rep. (2021) 48(1):691–699. 10.1007/s11033-020-06121-y.

Y. Zhou et al. Associations between genetic polymorphisms of TLRs and susceptibility to tuberculosis: A meta-analysis. Innate Immun. (2019) 26(2):75–83. 10.1177/1753425919862354.

