## Supplementary figures and images for "Identification of Blood miRNA Biomarkers in Systemic Tuberculosis through Metadata Analysis"

### supplementary Figure S5.png

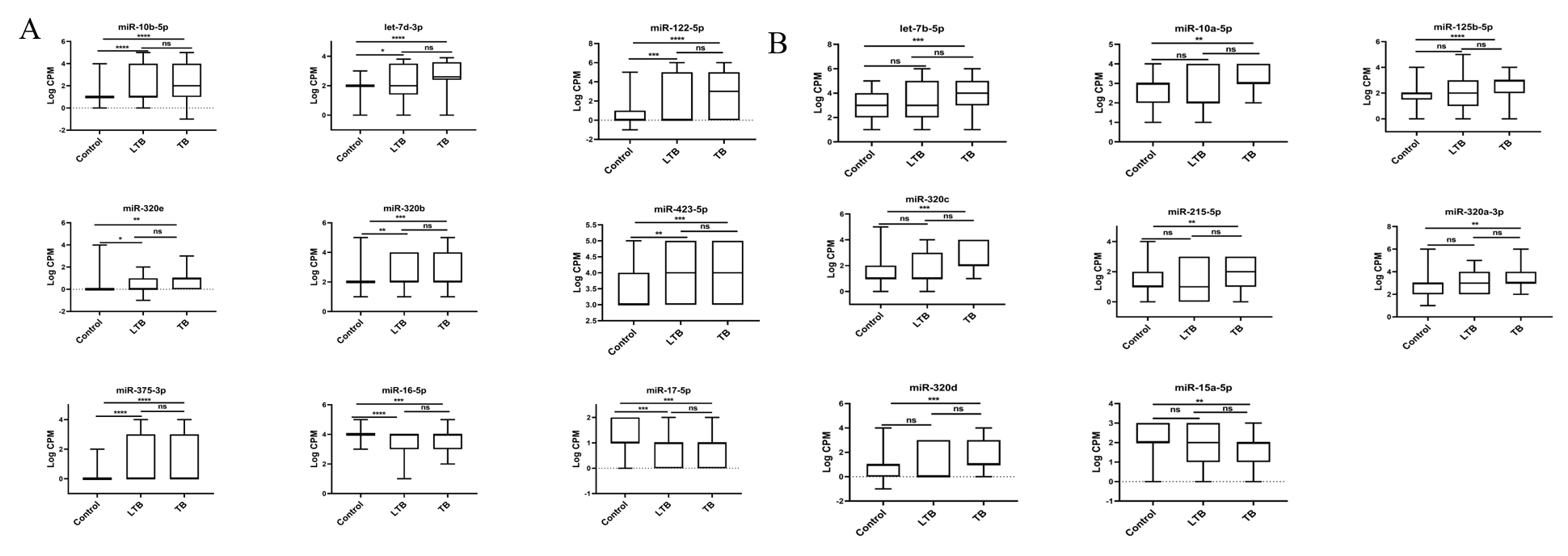

### supplementaryFigure S1.png

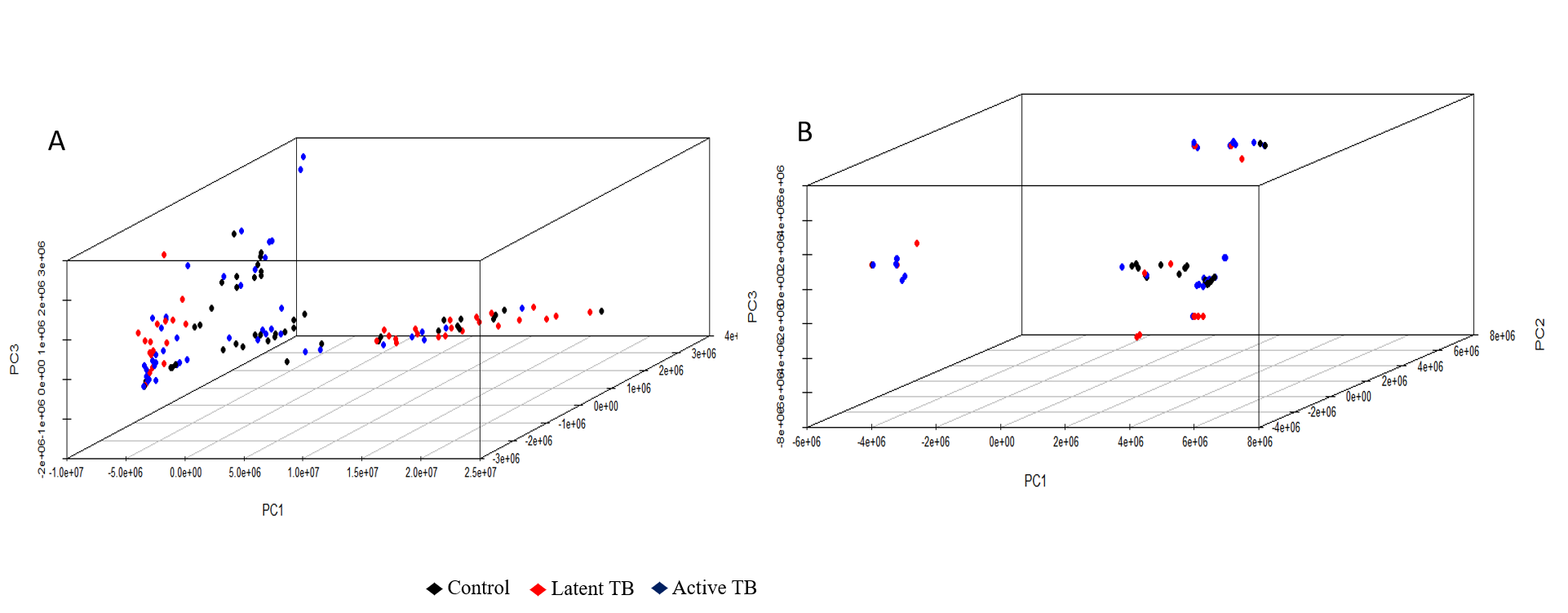

### supplementaryFigure S2.png

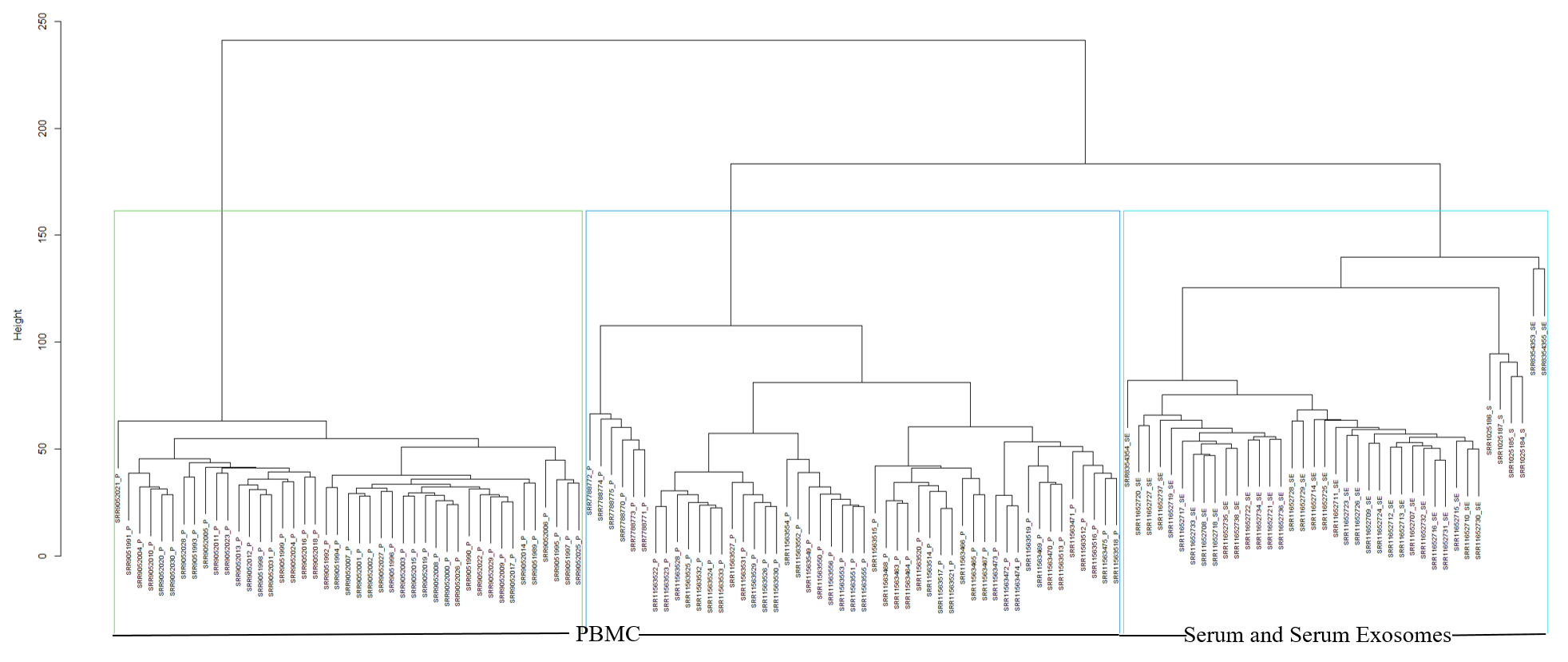

### supplementaryFigure S3.png

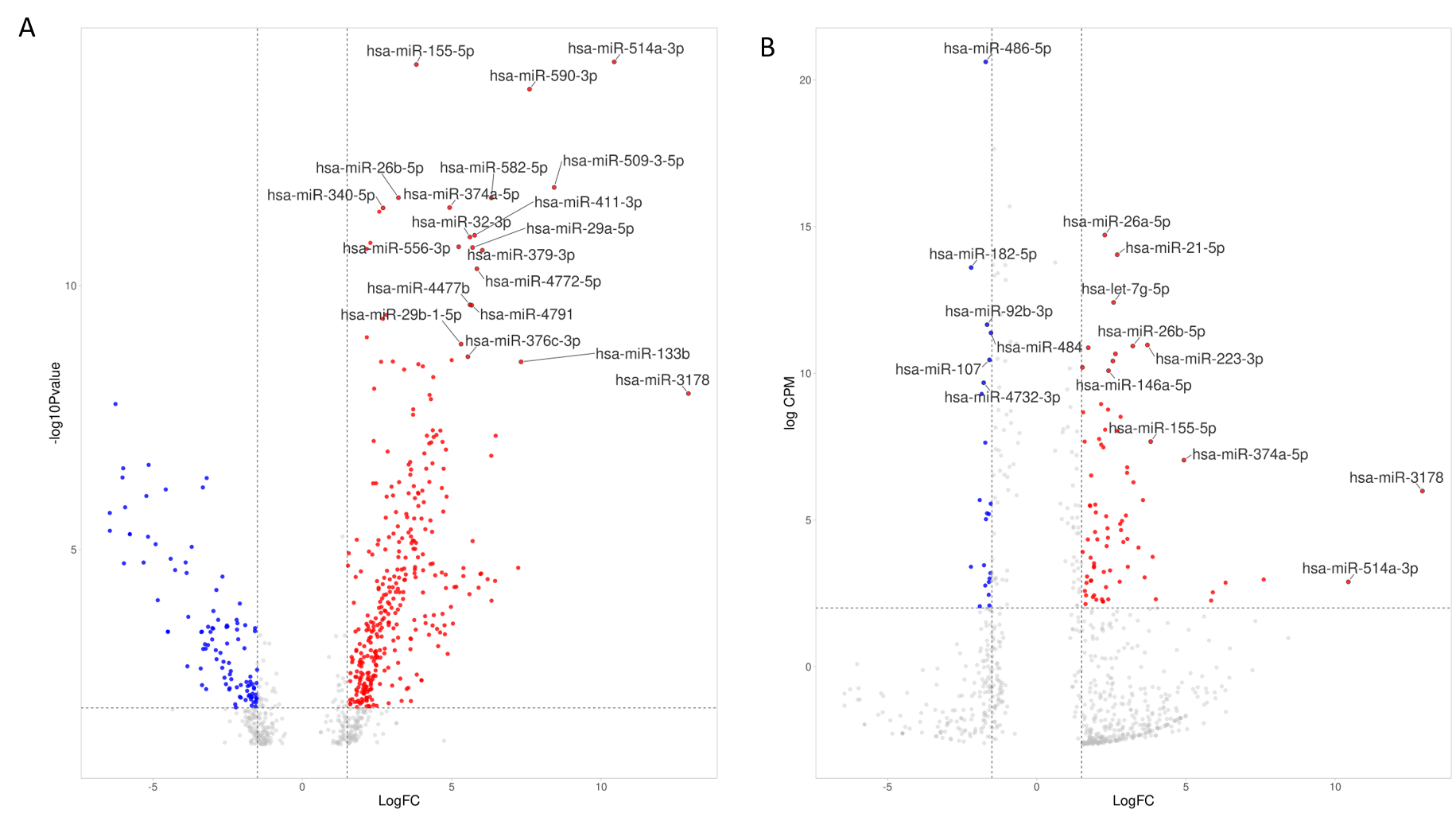

### supplementaryFigure S4.png

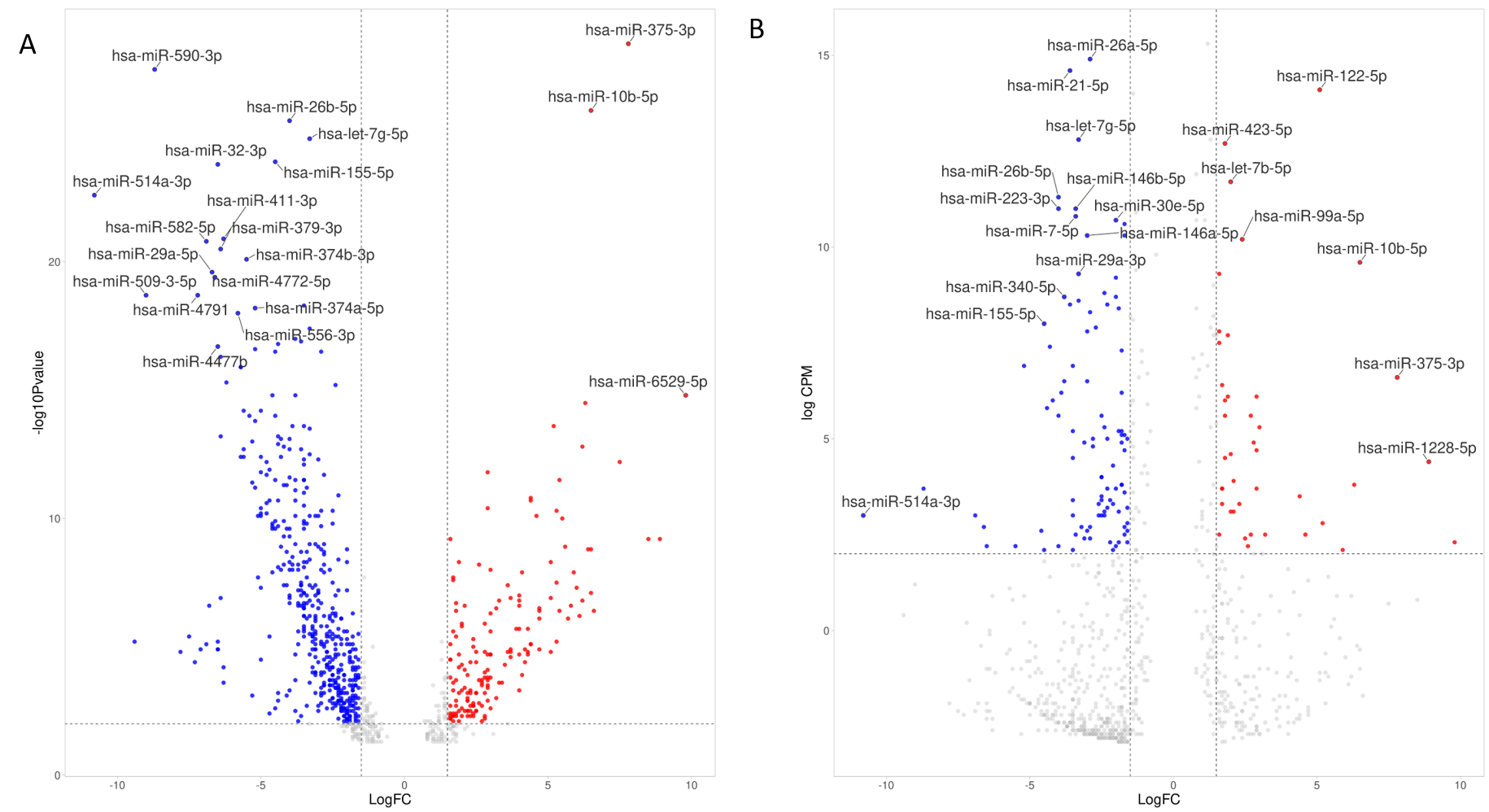

### supplementaryFigure S6.png

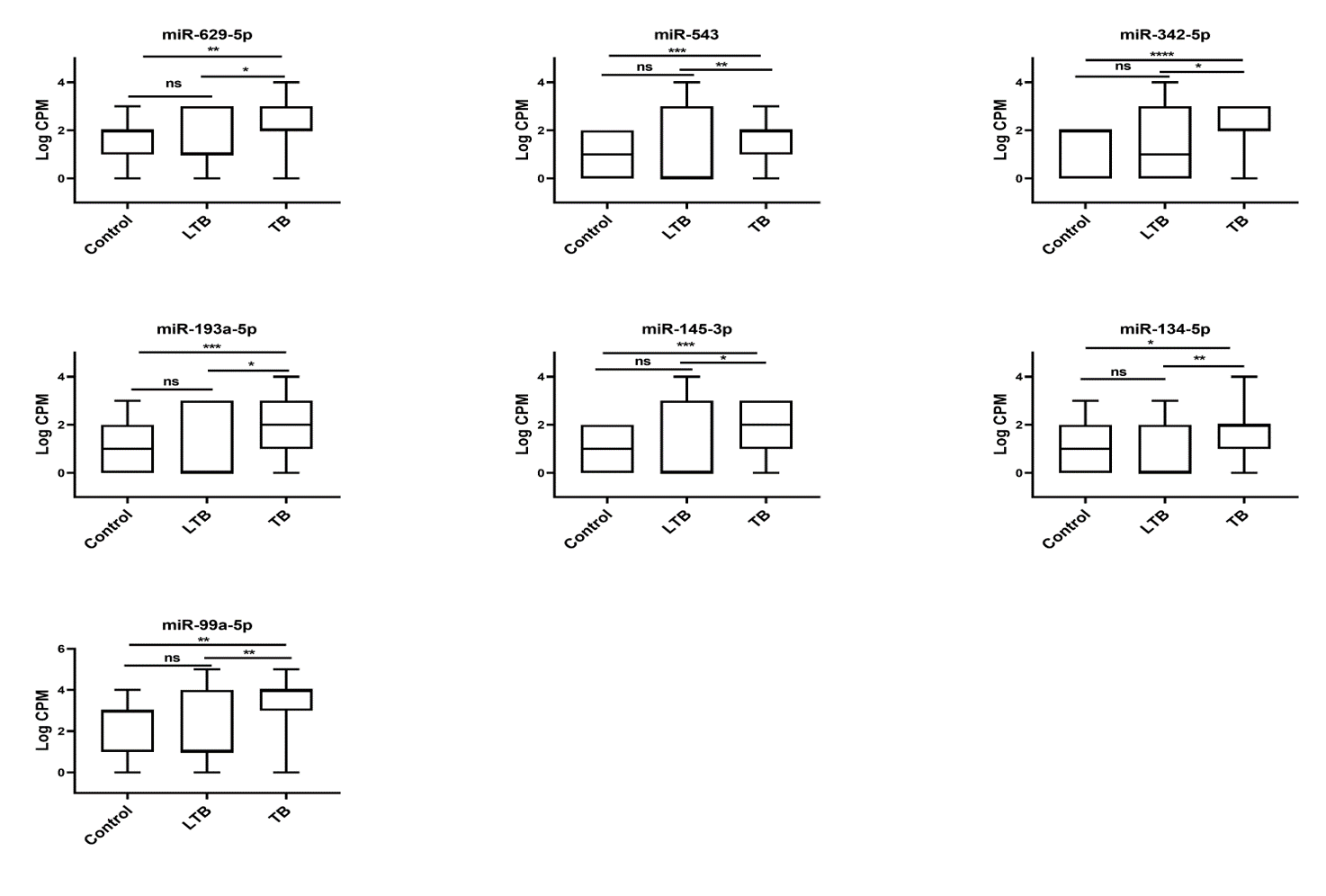

### supplementaryFigure S7.png

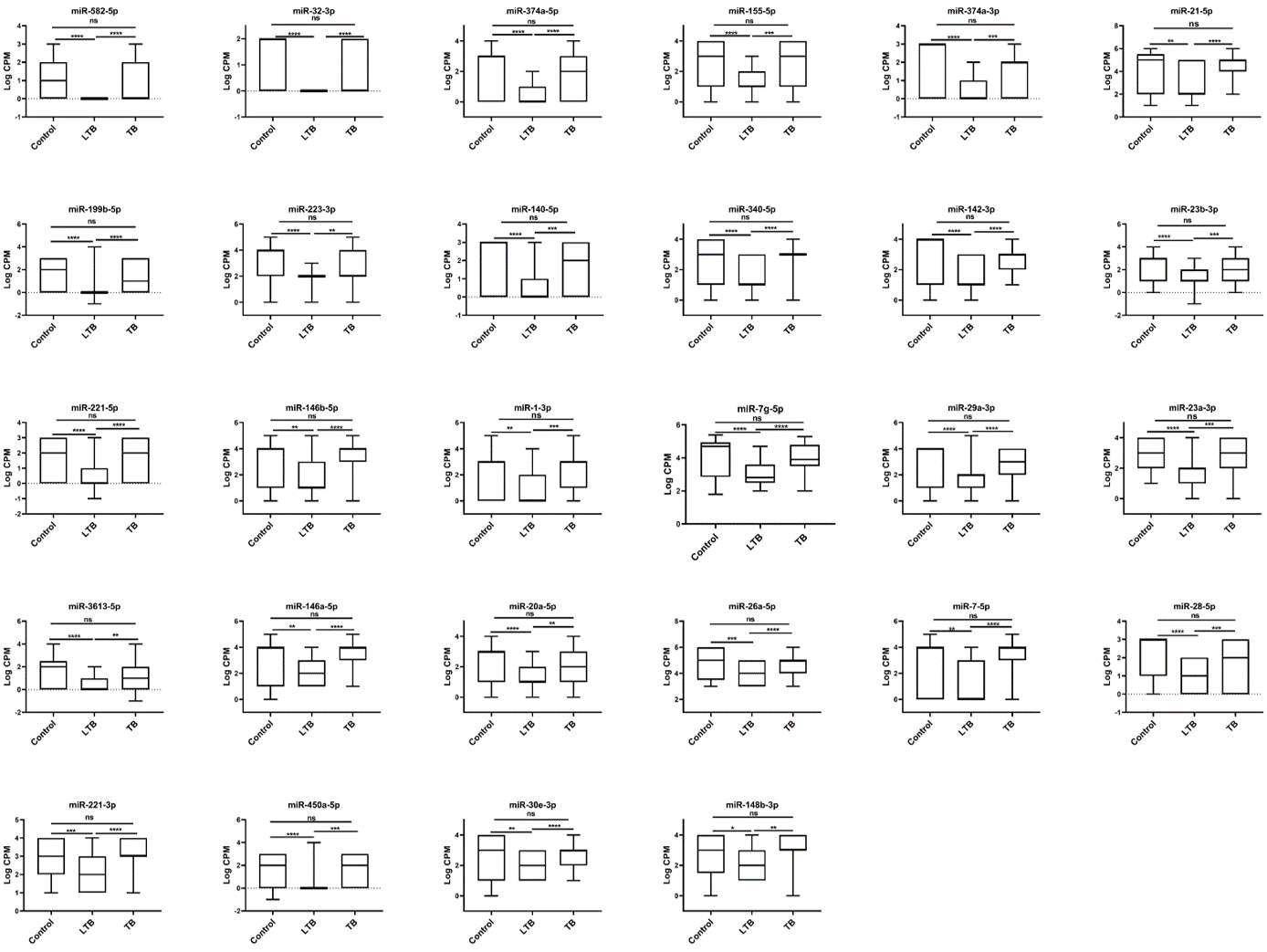

### supplementaryFigure S8.png

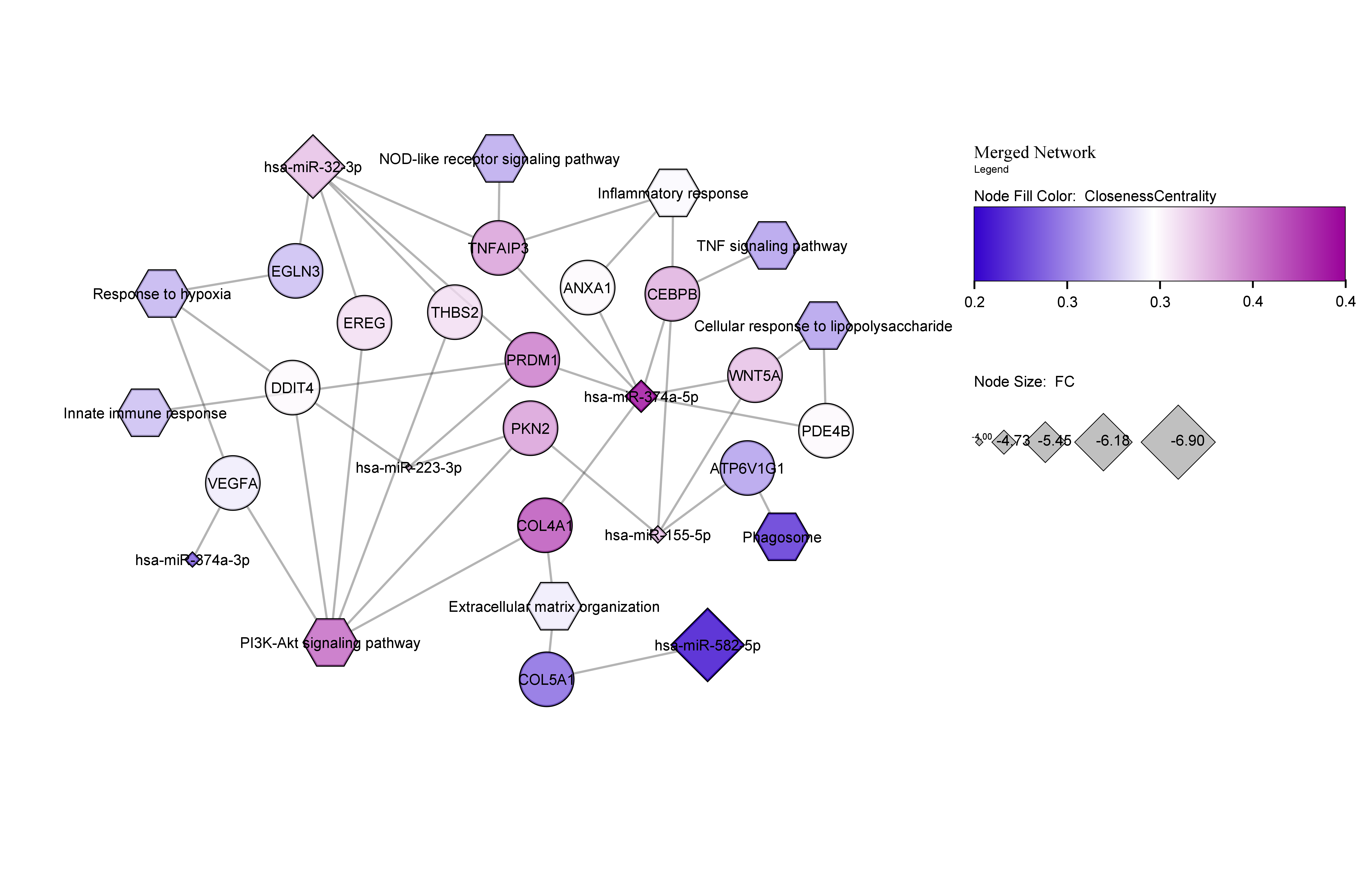

### supplementaryFigure S9.png

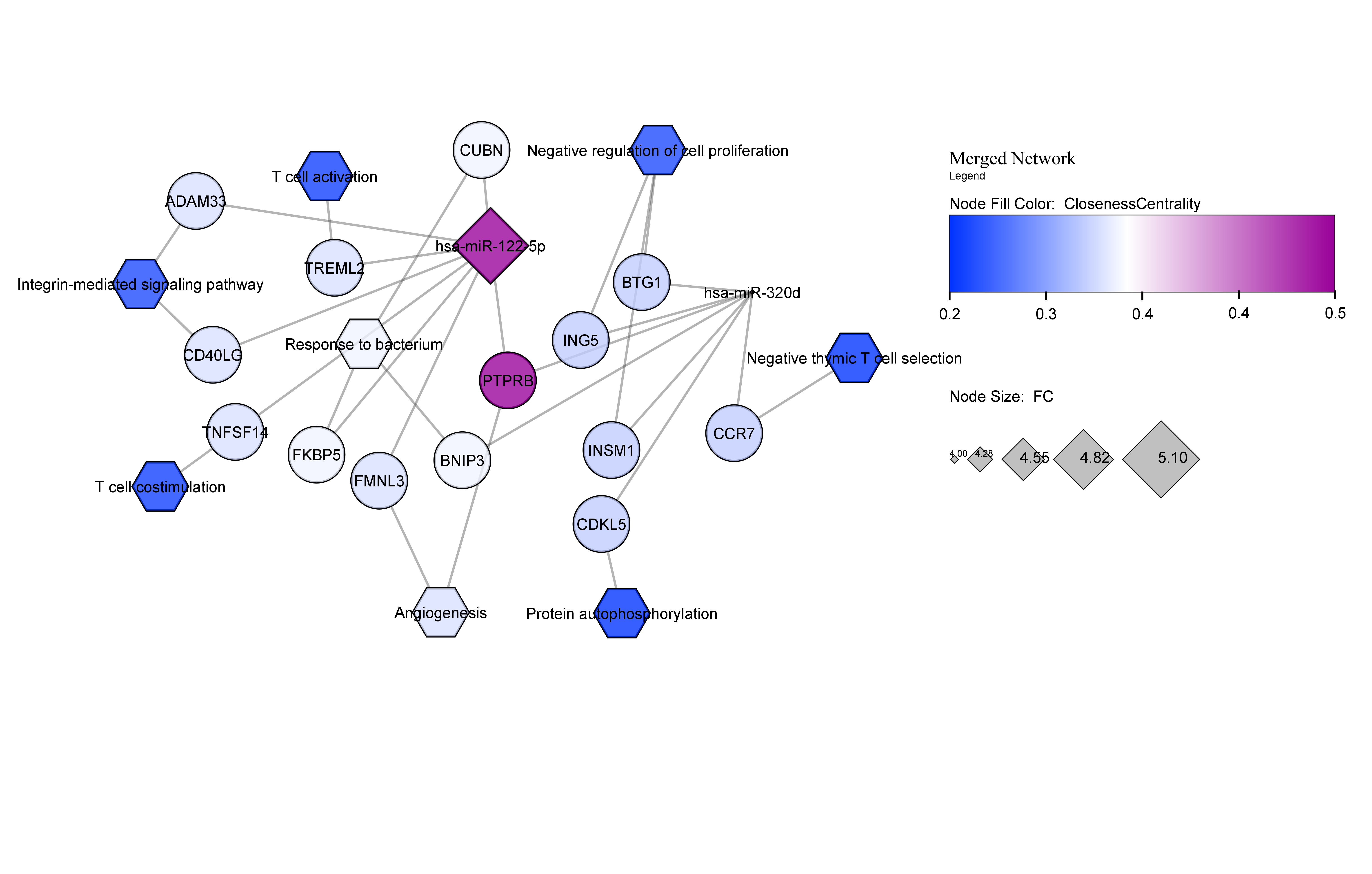
