## Supplementary material for "Identification of Blood miRNA Biomarkers in Systemic Tuberculosis through Metadata Analysis": Suplementary Tables: supplementary_tables 11Aug2023.docx.pdf

### Table S1. Meta- analysis sample details:

| S. NO. | Sample | Type# | Source* | NO. of reads (in Millions) | % Of Alignment | % Of Unmapped | Average mapped length | NO. of miRNAs |
|---|---|---|---|---|---|---|---|---|
| 1 | SRR1025184 | LTB1 | Serum | 8.0 | 83.18 | 16.82 | 18.5 | 161 |
| 2 | SRR1025185 | C1 | Serum | 8.5 | 81.46 | 18.54 | 18.4 | 127 |
| 3 | SRR1025186 | C2 | Serum | 8.7 | 83.5 | 16.5 | 18.9 | 215 |
| 4 | SRR1025187 | TB1 | Serum | 7.8 | 70.51 | 29.49 | 18.3 | 144 |
| 5 | SRR11563463 | C3 | PBMC | 21.0 | 99.72 | 0.28 | 24.2 | 957 |
| 6 | SRR11563464 | C4 | PBMC | 22.6 | 99.79 | 0.21 | 24.4 | 915 |
| 7 | SRR11563465 | C5 | PBMC | 22.5 | 99.73 | 0.27 | 24.2 | 924 |
| 8 | SRR11563466 | C6 | PBMC | 19.5 | 99.74 | 0.26 | 25.9 | 862 |
| 9 | SRR11563467 | C7 | PBMC | 22.6 | 99.71 | 0.29 | 24.8 | 915 |
| 10 | SRR11563468 | C8 | PBMC | 18.6 | 99.63 | 0.37 | 22.8 | 891 |
| 11 | SRR11563469 | C9 | PBMC | 20.7 | 99.7 | 0.3 | 24.6 | 803 |
| 12 | SRR11563470 | C10 | PBMC | 16.2 | 99.65 | 0.35 | 24.0 | 812 |
| 13 | SRR11563471 | C11 | PBMC | 15.0 | 99.08 | 0.92 | 22.8 | 762 |
| 14 | SRR11563472 | C12 | PBMC | 22.9 | 99.53 | 0.47 | 25.8 | 872 |
| 15 | SRR11563473 | C13 | PBMC | 20.1 | 99.69 | 0.31 | 26.0 | 819 |
| 16 | SRR11563474 | C14 | PBMC | 19.4 | 99.77 | 0.23 | 27.3 | 751 |
| 17 | SRR11563475 | C15 | PBMC | 16.7 | 99.36 | 0.64 | 26.9 | 735 |
| 18 | SRR11563512 | TB2 | PBMC | 20.6 | 99.36 | 0.64 | 25.9 | 835 |
| 19 | SRR11563513 | TB3 | PBMC | 19.5 | 99.63 | 0.37 | 24.9 | 844 |
| 20 | SRR11563514 | TB4 | PBMC | 24.4 | 99.62 | 0.38 | 22.8 | 926 |
| 21 | SRR11563515 | TB5 | PBMC | 25.2 | 99.18 | 0.82 | 24.1 | 911 |
| 22 | SRR11563516 | TB6 | PBMC | 22.7 | 99.43 | 0.57 | 25.4 | 838 |
| 23 | SRR11563517 | TB7 | PBMC | 19.6 | 99.63 | 0.37 | 23.5 | 879 |
| 24 | SRR11563518 | TB8 | PBMC | 20.2 | 99.73 | 0.27 | 25.1 | 806 |
| 25 | SRR11563519 | TB9 | PBMC | 20.8 | 99.81 | 0.19 | 32.1 | 630 |
| 26 | SRR11563520 | TB10 | PBMC | 20.3 | 99.47 | 0.53 | 23.4 | 936 |
| 27 | SRR11563521 | TB11 | PBMC | 19.0 | 99.72 | 0.28 | 23.4 | 883 |
| 28 | SRR11563522 | C16 | PBMC | 16.2 | 99.89 | 0.11 | 26.1 | 723 |
| 29 | SRR11563523 | C17 | PBMC | 16.7 | 99.91 | 0.09 | 26.3 | 757 |
| 30 | SRR11563524 | C18 | PBMC | 15.9 | 99.86 | 0.14 | 24.5 | 760 |
| 31 | SRR11563525 | C19 | PBMC | 17.2 | 99.89 | 0.11 | 29.3 | 699 |
| 32 | SRR11563526 | C20 | PBMC | 16.7 | 99.81 | 0.19 | 24.7 | 748 |
| 33 | SRR11563527 | C21 | PBMC | 15.8 | 99.87 | 0.13 | 27.3 | 687 |
| 34 | SRR11563528 | C22 | PBMC | 16.3 | 99.82 | 0.18 | 24.4 | 837 |
| 35 | SRR11563529 | C23 | PBMC | 15.2 | 99.87 | 0.13 | 25.4 | 692 |
| 36 | SRR11563530 | C24 | PBMC | 18.6 | 99.83 | 0.17 | 24.4 | 773 |
| 37 | SRR11563531 | C25 | PBMC | 15.4 | 99.91 | 0.09 | 28.3 | 639 |
| 38 | SRR11563532 | C26 | PBMC | 15.4 | 99.91 | 0.09 | 30.2 | 665 |
| 39 | SRR11563533 | C27 | PBMC | 14.2 | 99.9 | 0.1 | 27.5 | 716 |
| 40 | SRR11563549 | TB12 | PBMC | 12.2 | 99.84 | 0.16 | 27.5 | 754 |
| 41 | SRR11563550 | TB13 | PBMC | 15.7 | 99.81 | 0.19 | 27.4 | 749 |
| 42 | SRR11563551 | TB14 | PBMC | 16.3 | 99.78 | 0.22 | 27.4 | 788 |
| 43 | SRR11563552 | TB15 | PBMC | 16.5 | 99.92 | 0.08 | 29.7 | 660 |
| 44 | SRR11563553 | TB16 | PBMC | 14.9 | 99.88 | 0.12 | 25.5 | 730 |
| 45 | SRR11563554 | TB17 | PBMC | 2.0 | 99.82 | 0.18 | 26.9 | 420 |
| 46 | SRR11563555 | TB18 | PBMC | 15.2 | 99.88 | 0.12 | 26.8 | 772 |
| 47 | SRR11563556 | TB19 | PBMC | 16.2 | 99.79 | 0.21 | 23.8 | 835 |

| | | | | | | | | |
|---|---|---|---|---|---|---|---|---|
| 48 | SRR11652707 | LTB2 | Serum Exosomes | 13.0 | 90.58 | 9.42 | 19.3 | 297 |
| 49 | SRR11652708 | LTB3 | Serum Exosomes | 19.6 | 93.12 | 6.88 | 20.3 | 403 |
| 50 | SRR11652709 | LTB4 | Serum Exosomes | 19.3 | 95.2 | 4.8 | 20.9 | 429 |
| 51 | SRR11652710 | LTB5 | Serum Exosomes | 18.7 | 91.8 | 8.2 | 20.2 | 347 |
| 52 | SRR11652711 | LTB6 | Serum Exosomes | 17.2 | 93.64 | 6.36 | 20.6 | 397 |
| 53 | SRR11652712 | LTB7 | Serum Exosomes | 21.7 | 93.1 | 6.9 | 20.0 | 266 |
| 54 | SRR11652713 | LTB8 | Serum Exosomes | 25.4 | 91.08 | 8.92 | 20.3 | 239 |
| 55 | SRR11652714 | LTB9 | Serum Exosomes | 16.5 | 91.2 | 8.8 | 20.2 | 322 |
| 56 | SRR11652715 | LTB10 | Serum Exosomes | 11.8 | 94.02 | 5.98 | 20.6 | 436 |
| 57 | SRR11652716 | LTB11 | Serum Exosomes | 14.0 | 91.17 | 8.83 | 19.9 | 348 |
| 58 | SRR11652717 | LTB12 | Serum Exosomes | 13.2 | 94.29 | 5.71 | 20.7 | 240 |
| 59 | SRR11652718 | LTB13 | Serum Exosomes | 26.8 | 95.66 | 4.34 | 21.1 | 320 |
| 60 | SRR11652719 | LTB14 | Serum Exosomes | 16.3 | 93.47 | 6.53 | 21.2 | 247 |
| 61 | SRR11652720 | LTB15 | Serum Exosomes | 24.1 | 92.75 | 7.25 | 20.0 | 217 |
| 62 | SRR11652721 | LTB16 | Serum Exosomes | 18.1 | 90.46 | 9.54 | 19.4 | 214 |
| 63 | SRR11652722 | LTB17 | Serum Exosomes | 27.7 | 92.94 | 7.06 | 19.5 | 221 |
| 64 | SRR11652723 | TB20 | Serum Exosomes | 11.3 | 92.11 | 7.89 | 20.2 | 370 |
| 65 | SRR11652724 | TB21 | Serum Exosomes | 15.5 | 93.1 | 6.9 | 20.6 | 376 |
| 66 | SRR11652725 | TB22 | Serum Exosomes | 18.7 | 90.09 | 9.91 | 19.8 | 230 |
| 67 | SRR11652726 | TB23 | Serum Exosomes | 9.3 | 86.79 | 13.21 | 18.5 | 202 |
| 68 | SRR11652727 | TB24 | Serum Exosomes | 15.9 | 87.83 | 12.17 | 19.1 | 207 |
| 69 | SRR11652728 | TB25 | Serum Exosomes | 15.5 | 90.23 | 9.77 | 20.2 | 241 |
| 70 | SRR11652729 | TB26 | Serum Exosomes | 14.1 | 87.6 | 12.4 | 18.2 | 146 |
| 71 | SRR11652730 | TB27 | Serum Exosomes | 8.9 | 90.19 | 9.81 | 20.0 | 186 |
| 72 | SRR11652731 | TB28 | Serum Exosomes | 14.5 | 86.07 | 13.93 | 18.3 | 168 |
| 73 | SRR11652732 | TB29 | Serum Exosomes | 10.4 | 87.98 | 12.02 | 19.6 | 167 |
| 74 | SRR11652733 | TB30 | Serum Exosomes | 24.2 | 92.96 | 7.04 | 20.8 | 187 |
| 75 | SRR11652734 | TB31 | Serum Exosomes | 13.3 | 94.06 | 5.94 | 20.8 | 167 |

| | | | | | | | | |
|---|---|---|---|---|---|---|---|---|
| 76 | SRR11652735 | TB32 | Serum Exosomes | 20.8 | 94.57 | 5.43 | 21.3 | 294 |
| 77 | SRR11652736 | TB33 | Serum Exosomes | 8.0 | 92.22 | 7.78 | 20.7 | 241 |
| 78 | SRR11652737 | TB34 | Serum Exosomes | 13.4 | 94.8 | 5.2 | 21.4 | 299 |
| 79 | SRR11652738 | TB35 | Serum Exosomes | 3.7 | 91.26 | 8.74 | 20.2 | 216 |
| 80 | SRR7788770 | TB36 | PBMC | 8.7 | 100 | 0 | 23.7 | 640 |
| 81 | SRR7788771 | TB37 | PBMC | 7.8 | 99.99 | 0.01 | 24.1 | 626 |
| 82 | SRR7788772 | TB38 | PBMC | 6.9 | 100 | 0 | 24.0 | 601 |
| 83 | SRR7788773 | C28 | PBMC | 6.9 | 99.99 | 0.01 | 24.6 | 629 |
| 84 | SRR7788774 | C29 | PBMC | 7.7 | 100 | 0 | 23.4 | 518 |
| 85 | SRR7788775 | C30 | PBMC | 8.6 | 100 | 0 | 23.0 | 597 |
| 86 | SRR8354353 | C31 | Serum exosomes | 12.8 | 99.37 | 0.63 | 30.9 | 115 |
| 87 | SRR8354354 | LTB18 | Serum exosomes | 1.2 | 94.91 | 5.09 | 27.9 | 176 |
| 88 | SRR8354355 | TB39 | Serum exosomes | 15.8 | 88.92 | 11.08 | 18.1 | 214 |
| 89 | SRR9051989 | TB40 | PBMC | 10.9 | 100 | 0 | 24.3 | 443 |
| 90 | SRR9051990 | TB41 | PBMC | 22.9 | 100 | 0 | 24.3 | 511 |
| 91 | SRR9051991 | TB42 | PBMC | 12.5 | 100 | 0 | 24.7 | 407 |
| 92 | SRR9051992 | TB43 | PBMC | 16.2 | 100 | 0 | 24.4 | 505 |
| 93 | SRR9051993 | TB44 | PBMC | 13.4 | 100 | 0 | 24.9 | 353 |
| 94 | SRR9051994 | TB45 | PBMC | 10.2 | 100 | 0 | 24.3 | 466 |
| 95 | SRR9051995 | TB46 | PBMC | 15.0 | 100 | 0 | 24.7 | 444 |
| 96 | SRR9051996 | TB47 | PBMC | 18.1 | 100 | 0 | 24.1 | 497 |
| 97 | SRR9051997 | C32 | PBMC | 16.0 | 100 | 0 | 24.7 | 428 |
| 98 | SRR9051998 | C33 | PBMC | 16.0 | 100 | 0 | 24.5 | 459 |
| 99 | SRR9051999 | C34 | PBMC | 15.9 | 100 | 0 | 24.7 | 389 |
| 100 | SRR9052000 | C35 | PBMC | 19.3 | 100 | 0 | 24.2 | 545 |
| 101 | SRR9052001 | C36 | PBMC | 12.9 | 100 | 0 | 24.0 | 502 |
| 102 | SRR9052002 | C37 | PBMC | 14.7 | 100 | 0 | 24.6 | 533 |
| 103 | SRR9052003 | C38 | PBMC | 18.7 | 100 | 0 | 24.0 | 539 |
| 104 | SRR9052004 | C39 | PBMC | 13.2 | 100 | 0 | 24.4 | 435 |
| 105 | SRR9052005 | C40 | PBMC | 7.1 | 100 | 0 | 24.8 | 340 |
| 106 | SRR9052006 | C41 | PBMC | 21.4 | 100 | 0 | 25.3 | 361 |
| 107 | SRR9052007 | C42 | PBMC | 18.9 | 100 | 0 | 24.4 | 486 |
| 108 | SRR9052008 | C43 | PBMC | 22.3 | 100 | 0 | 24.4 | 577 |
| 109 | SRR9052009 | C44 | PBMC | 20.8 | 100 | 0 | 24.0 | 550 |
| 110 | SRR9052010 | C45 | PBMC | 17.5 | 100 | 0 | 24.3 | 488 |
| 111 | SRR9052011 | LTB19 | PBMC | 20.0 | 100 | 0 | 24.6 | 534 |
| 112 | SRR9052012 | LTB20 | PBMC | 19.4 | 100 | 0 | 25.0 | 400 |
| 113 | SRR9052013 | LTB21 | PBMC | 21.0 | 100 | 0 | 24.9 | 412 |
| 114 | SRR9052014 | LTB22 | PBMC | 16.7 | 100 | 0 | 24.3 | 463 |
| 115 | SRR9052015 | LTB23 | PBMC | 16.6 | 100 | 0 | 23.9 | 549 |
| 116 | SRR9052016 | LTB24 | PBMC | 14.1 | 100 | 0 | 24.7 | 437 |
| 117 | SRR9052017 | LTB25 | PBMC | 21.8 | 100 | 0 | 24.2 | 526 |
| 118 | SRR9052018 | LTB26 | PBMC | 14.3 | 100 | 0 | 24.9 | 409 |
| 119 | SRR9052019 | LTB27 | PBMC | 16.3 | 100 | 0 | 24.1 | 494 |
| 120 | SRR9052020 | LTB28 | PBMC | 14.9 | 100 | 0 | 24.5 | 437 |
| 121 | SRR9052021 | LTB29 | PBMC | 16.1 | 100 | 0 | 24.8 | 375 |
| 122 | SRR9052022 | LTB30 | PBMC | 15.9 | 100 | 0 | 24.0 | 541 |
| 123 | SRR9052023 | LTB31 | PBMC | 19.6 | 100 | 0 | 24.9 | 416 |
| 124 | SRR9052024 | LTB32 | PBMC | 17.4 | 100 | 0 | 24.7 | 421 |

| | | | | | | | | |
|---|---|---|---|---|---|---|---|---|
| 125 | SRR9052025 | LTB33 | PBMC | 13.4 | 100 | 0 | 24.5 | 452 |
| 126 | SRR9052026 | LTB34 | PBMC | 19.2 | 100 | 0 | 24.2 | 541 |
| 127 | SRR9052027 | LTB35 | PBMC | 20.8 | 100 | 0 | 24.2 | 573 |
| 128 | SRR9052028 | LTB36 | PBMC | 14.4 | 100 | 0 | 24.4 | 397 |
| 129 | SRR9052029 | LTB37 | PBMC | 22.5 | 100 | 0 | 24.5 | 517 |
| 130 | SRR9052030 | LTB38 | PBMC | 18.4 | 100 | 0 | 24.3 | 486 |
| 131 | SRR9052031 | LTB39 | PBMC | 13.0 | 100 | 0 | 25.0 | 405 |

#C1 to C45 – Control, TB1 to TB47 - Active TB, and LTB1 to LTB39 - Latent TB

*PBMC- Peripheral Blood Mononuclear Cell

### Table S2. TB-specific miRNAs in blood:

| S.NO. | miRNA | logFC | Log CPM | -Log10P-Value |
|---|---|---|---|---|
| **Up regulated** | | | | |
| 1 | hsa-miR-10b-5p | 7.1 | 10.3 | 25.8 |
| 2 | hsa-miR-375-3p | 6.9 | 6.3 | 20.9 |
| 3 | miR-122-5p | 5.1 | 14.3 | 5.3 |
| 4 | hsa-miR-320b | 2.8 | 10.3 | 7.7 |
| 5 | hsa-let-7d-3p | 2.6 | 8.8 | 10.6 |
| 6 | hsa-miR-423-5p | 2.3 | 13.5 | 7.3 |
| 7 | miR-320e | 1.8 | 3.2 | 2.0 |
| **Down regulated** | | | | |
| 1 | hsa-miR-16-5p | -1.2 | 13.6 | 2.1 |
| 2 | hsa-miR-17-3p | -1.2 | 5.0 | 2.0 |

Differentially expressed miRNAs in LTB patients (Expression levels were same in both active TB and controls) compared to healthy controls. The miRNAs with the absolute fold change >1.2 and adj.p <0.05 and log CPM greater than 2

**Table S3. Dysregulated miRNAs in Active TB compared to healthy controls:**

| S.NO. | miRNA | logFC | Log CPM | -Log10P-Value |
|---|---|---|---|---|
| **Up regulated** | | | | |
| 1 | hsa-miR-320d | 4.0 | 7.7 | 10.5 |
| 2 | hsa-miR-320c | 3.0 | 9.0 | 7.6 |
| 3 | hsa-let-7b-5p | 2.5 | 12.5 | 8.6 |
| 4 | hsa-miR-320a-3p | 2.5 | 12.0 | 9.2 |
| 5 | hsa-miR-215-5p | 2.1 | 6.3 | 7.0 |
| 6 | hsa-miR-125b-5p | 2 | 7.1 | 11.0 |
| 7 | hsa-miR-10a-5p | 1.6 | 9.9 | 7.0 |
| **Down regulated** | | | | |
| 1 | hsa-miR-15a-5p | -1.6 | 9.2 | 2.4 |

Differentially expressed miRNAs in LTB patients (Expression levels were same in both active TB and controls) compared to healthy controls. The miRNAs with the absolute fold change >1.2 and adj.p <0.05 and log CPM greater than 2

**Table S4. Dysregulated miRNAs in TB progression:**

| S.NO. | miRNA | logFC | logCPM | -Log10P-Value |
|---|---|---|---|---|
| 1 | hsa-miR-193a-5p | 3.9 | 6.3 | 16.7 |
| 2 | hsa-miR-629-5p | 3.1 | 7.5 | 13.2 |
| 3 | hsa-miR-342-5p | 2.9 | 7 | 11.7 |
| 4 | hsa-miR-134-5p | 2.9 | 5.6 | 9.0 |
| 5 | hsa-miR-145-3p | 2.7 | 5.8 | 8.7 |
| 6 | hsa-miR-543 | 2.6 | 5.6 | 7.2 |
| 7 | hsa-miR-99a-5p | 2.6 | 11.2 | 7.3 |

Disease progressive miRNAs. The miRNAs with the absolute fold change >1.2 and adj.p <0.05 and log CPM greater than 2.

### Table S5. LTB-specific miRNAs in blood:

| S.NO. | miRNA | logFC | Log CPM | -Log10P-Value |
|---|---|---|---|---|
| 1 | hsa-miR-582-5p | -6.9 | 3.0 | 20.8 |
| 2 | hsa-miR-32-3p | -6.5 | 2.2 | 23.8 |
| 3 | hsa-miR-374a-5p | -5.2 | 6.9 | 18.2 |
| 4 | hsa-miR-155-5p | -4.5 | 8.0 | 23.9 |
| 5 | hsa-miR-374a-3p | -4.4 | 5.8 | 16.8 |
| 6 | hsa-miR-199b-5p | -4.2 | 6.0 | 12.8 |
| 7 | hsa-miR-223-3p | -4.0 | 11 | 12.7 |
| 8 | hsa-miR-140-5p | -4.0 | 5.6 | 13.1 |
| 9 | hsa-miR-340-5p | -3.8 | 8.7 | 17.0 |
| 10 | hsa-miR-142-3p | -3.6 | 8.5 | 16.9 |
| 11 | hsa-miR-21-5p | -3.6 | 14.6 | 12.7 |
| 12 | hsa-miR-23b-3p | -3.5 | 6.9 | 18.3 |
| 13 | hsa-miR-221-5p | -3.5 | 5.2 | 11.0 |
| 14 | hsa-miR-146b-5p | -3.4 | 11.0 | 11.2 |
| 15 | hsa-miR-7-5p | -3.4 | 10.8 | 6.7 |
| 16 | hsa-miR-7g-5p | -3.3 | 12.8 | 24.8 |
| 17 | hsa-miR-29a-3p | -3.3 | 9.3 | 13.5 |
| 18 | hsa-miR-23a-3p | -3.3 | 8.6 | 17.4 |
| 19 | hsa-miR-3613-5p | -3.1 | 4.9 | 10.1 |
| 20 | hsa-miR-146a-5p | -3.0 | 10.3 | 10.10 |
| 21 | hsa-miR-20a-5p | -3.0 | 7.8 | 12.3 |
| 22 | hsa-miR-26a-5p | -2.9 | 14.9 | 16.5 |
| 23 | hsa-miR-1-3p | -2.7 | 7.9 | 2.6 |
| 24 | hsa-miR-28-5p | -2.5 | 5.6 | 10.3 |
| 25 | hsa-miR-221-3p | -2.4 | 8.8 | 15.2 |
| 26 | hsa-miR-450a-5p | -2.4 | 5.3 | 4.7 |
| 27 | hsa-miR-30e-3p | -2.3 | 8.5 | 10.9 |
| 28 | hsa-miR-148b-3p | -2.0 | 9.2 | 7.2 |

Differentially expressed miRNAs in LTB patients (Expression levels were same in both active TB and controls) compared to healthy controls. The miRNAs with the absolute fold change >1.2 and adj.p <0.05 and log CPM greater than 2.

**Table S6. Biological pathways upregulated in TB (Downregulated miRNAs):**

| S.NO. | GOTERM_BP_DIRECT | Genes | P-Value | FDR |
|---|---|---|---|---|
| 1 | *Inflammatory response | 24 | 5.30E-07 | 9.40E-04 |
| 2 | Defense response to virus | 19 | 7.20E-07 | 9.40E-04 |
| 3 | Cell adhesion | 28 | 1.20E-05 | 1.00E-02 |
| 4 | Apoptotic process | 29 | 2.20E-05 | 1.10E-02 |
| 5 | *Cellular response to interferon-gamma | 9 | 2.20E-05 | 1.10E-02 |
| 6 | Endodermal cell differentiation | 7 | 2.50E-05 | 1.10E-02 |
| 7 | Immune response | 25 | 3.60E-05 | 1.30E-02 |
| 8 | *Response to hypoxia | 14 | 5.70E-05 | 1.90E-02 |
| 9 | Positive regulation of apoptotic process | 19 | 7.80E-05 | 2.30E-02 |
| 10 | *Extracellular matrix organization | 13 | 9.30E-05 | 2.40E-02 |
| 11 | *Cellular response to lipopolysaccharide | 12 | 1.00E-04 | 2.50E-02 |
| 12 | *Innate immune response | 25 | 2.90E-04 | 5.80E-02 |
| 13 | Positive regulation of GTPase activity | 13 | 3.30E-04 | 5.80E-02 |
| 14 | *Leukocyte cell-cell adhesion | 6 | 3.50E-04 | 5.80E-02 |
| 15 | *Positive regulation of nitric oxide biosynthetic process | 6 | 3.50E-04 | 5.80E-02 |

*Interested pathways associated with tuberculosis pathogenesis selected for further analysis

**Table S7. KEGG signaling pathways upregulated in TB (Downregulated miRNAs):**

| S.NO. | KEGG pathways | Genes | P-Value |
|---|---|---|---|
| 1 | Malaria | 10 | 4.50E-06 |
| 2 | AGE-RAGE signaling pathway in diabetic complications | 12 | 5.20E-05 |
| 3 | Lipid and atherosclerosis | 16 | 4.60E-04 |
| 4 | *TNF signaling pathway | 8 | 2.50E-03 |
| 5 | Proteoglycans in cancer | 14 | 2.60E-03 |
| 6 | *Phagosome | 10 | 6.10E-03 |
| 7 | Yersinia infection | 10 | 9.20E-03 |
| 8 | Rheumatoid arthritis | 8 | 1.00E-02 |
| 9 | *PI3K-Akt signaling pathway | 18 | 1.10E-02 |
| 10 | Protein digestion and absorption | 8 | 1.70E-02 |
| 11 | *NOD-like receptor signaling pathway | 10 | 2.10E-02 |
| 12 | ECM-receptor interaction | 7 | 2.70E-02 |
| 13 | Transcriptional misregulation in cancer | 11 | 2.90E-02 |
| 14 | Prolactin signaling pathway | 6 | 3.50E-02 |
| 15 | Pertussis | 6 | 4.70E-02 |

*Interested pathways associated with tuberculosis pathogenesis selected for further analysis

**Table S8. Biological pathways downregulated in TB (Upregulated miRNAs):**

| S.NO. | GOTERM_BP_DIRECT | Genes | P-Value |
|---|---|---|---|
| 1 | Activation of GTPase activity | 7 | 8.70E-04 |
| 2 | *Protein autophosphorylation | 8 | 1.00E-03 |
| 3 | *Positive regulation of angiogenesis | 7 | 3.10E-03 |
| 4 | *B cell activation | 4 | 3.70E-03 |
| 5 | *Negative thymic T cell selection | 3 | 3.90E-03 |
| 6 | Negulation of transcription, DNA-templated | 18 | 4.80E-03 |
| 7 | *T cell costimulation | 4 | 5.30E-03 |
| 8 | Regulation of transcription from RNA polymerase II promoter | 26 | 7.10E-03 |
| 9 | *Negative regulation of cell proliferation | 12 | 7.20E-03 |
| 10 | Cell adhesion | 12 | 9.00E-03 |
| 11 | *Response to growth factor | 3 | 9.30E-03 |
| 12 | Action potential | 3 | 9.30E-03 |
| 13 | *T cell activation | 4 | 9.80E-03 |
| 14 | Immune response | 11 | 1.10E-02 |
| 15 | *Integrin-mediated signaling pathway | 5 | 1.50E-02 |
| 16 | Negative regulation of apoptotic process | 11 | 1.60E-02 |
| 17 | Apoptotic process | 12 | 1.70E-02 |
| 18 | *Intracellular signal transduction | 11 | 1.70E-02 |
| 19 | *CD8-positive, alpha-beta T cell lineage commitment | 2 | 1.70E-02 |
| 20 | Regulation of endodeoxyribonuclease activity | 2 | 1.70E-02 |
| 21 | Cell aging | 3 | 2.00E-02 |
| 22 | Neuron migration | 5 | 2.00E-02 |
| 23 | *Response to bacterium | 7 | 2.10E-02 |
| 24 | *Angiogenesis | 7 | 2.30E-02 |
| 25 | Positive regulation of apoptotic process | 8 | 2.40E-02 |

*Interested pathways associated with tuberculosis pathogenesis selected for further analysis

**Table S9. KEGG signaling pathways downregulated in TB (Upregulated miRNAs):**

| S.NO. | KEGG pathways | Genes | P-Value |
|---|---|---|---|
| 1 | *NF-kappa B signaling pathway | 6 | 2.70E-04 |
| 2 | Herpes simplex virus 1 infection | 12 | 5.80E-03 |
| 3 | *Allograft rejection | 3 | 4.80E-02 |
| 4 | Small cell lung cancer | 4 | 5.40E-02 |
| 5 | Cell adhesion molecules | 5 | 5.80E-02 |
| 6 | Ras signaling pathway | 6 | 6.80E-02 |
| 7 | Calcium signaling pathway | 6 | 7.20E-02 |
| 8 | T cell receptor signaling pathway | 4 | 7.20E-02 |
| 9 | Autoimmune thyroid disease | 3 | 8.90E-02 |

*Interested pathways associated with tuberculosis pathogenesis selected for further analysis

**Table S10. Downregulated miRNAs and their target genes pathways**

| S.NO. | miRNAs and their Targets | KEGG pathways |
|---|---|---|
| 1 | let-7g-5p [IL6], miR-146a-5p [CCL5], miR-146b-5p [CCL5], miR-155-5p [CEBPB], miR-17-5p [CREB1], miR-20a-5p [CREB1], miR-23a-3p [TNFAIP3], miR-23b-3p [ TNFAIP3], miR-26a-5p [CREB1], miR-29a-3p [TNFAIP3, PIK3CB], miR-30e-3p [CREB1], miR-32-3p [BCL3, TNFAIP3], miR-374a-5p [CEBPB, SELE, TNFAIP3] | TNF signaling pathway |
| 2 | let-7g-5p [ATP6V1G1, FCAR, MSR1, OLR1], miR-148b-3p [CTSL, ITGA5, TFRC], miR-155-5p [ATP6V1G1, TLR6], miR-17-5p [MSR1, THBS2], miR-20a-5p [MSR1, THBS2], miR-21-5p [OLR1], miR-26a-5p [FCAR, ITGA5], miR-29a-3p [THBS2], miR-30e-3p [ITGA5, MSR1], miR-32-3p [THBS2], miR-7-5p [FCAR, TFRC, TLR4] | Phagosome |
| 3 | miR-148b-3p [COL4A1, COL6A3, ITGA5, KIT], miR-23b-3p [COL4A1], miR-23a-3p [COL4A1], miR-29a-3p [COL4A1, COL6A3, PIK3CB, THBS2, VEGFA], miR-20a-5p [COL4A1, CREB1, EREG, OSM, PKN2, THBS2, VEGFA], let-7g-5p [COL4A1, IL6, PKN2], miR-1-3p [FN1, VEGFA], miR-155-5p [PKN2], miR-15a-5p [VEGFA], miR-16-5p [VEGFA], miR-17-5p [COL4A1, CREB1, EREG, OSM, PKN2, THBS2, VEGFA], miR-221-3p [DDIT4, KIT], miR-223-3p [DDIT4, LAMB1, PKN2], miR-26a-5p [CREB1, EREG, ITGA5, LPAR3], miR-30e-3p [CREB1, ITGA5], miR-32-3p [EREG, THBS2], miR-374a-3p [VEGFA], miR-374a-5p [COL4A1], miR-582-5p [PIK3AP1], miR-7-5p [DDIT4, TLR4] | PI3K-Akt signaling pathway |

| | | |
|---|---|---|
| 4 | let-7g-5p [ IL6, PKN2], miR-1-3p [NAMPT], miR-146a-5p [CCL5], miR-146b-5p [CCL5], miR-148b-3p[PANX1], miR-155-5p [PKN2], miR-17-5p [GBP3, PKN2], miR-20a-5p [GBP3, PKN2], miR-223-3p [PKN2], miR-23a-3p [GBP3, TNFAIP3, TXN], miR-23b-3p [GBP3, TNFAIP3, TXN], miR-26a-5p [GBP1, NAMPT], miR-29a-3p [TNFAIP3], miR-30e-3p [GBP1], miR-32-3p [TNFAIP3], miR-374a-5p [TNFAIP3], miR-7-5p [TLR4] | NOD-like receptor signaling pathway |
| **S.NO.** | **miRNAs and their Targets** | **GO pathways** |
| 1 | let-7g-5p [CCL7, IL6, OLR1], miR-146a-5p [CCL5, NOX4], miR-146b-5p [CCL5, NOX4], miR-148b-3p [CHST1, KIT], miR-155-5p [CEBPB, TLR6], miR-15a-5p [CD40, IRAK2, TLR1], miR-16-5p [CD40, IRAK2, TLR1], miR-21-5p [ANXA1, OLR1], miR-221-3p [KIT], miR-223-3p [LACC1], miR-23a-3p [CCL7, PROK2, TNFAIP3, TPST1], miR-23b-3p [CCL7, PROK2, TNFAIP3, TPST1], miR-26a-5p [ADM, SELP], miR-29a-3p [F11R, PTX3, TNFAIP3], miR-30e-3p [ANXA1, APOL3, CCL7], miR-32-3p [TNFAIP3], miR-3613-5p [F11R], miR-374a-5p [ANXA1, CEBPB, SELE, TNFAIP3], miR-7-5p [TLR4] | Inflammatory response |
| 2 | let-7g-5p [CCL7, FCAR], miR-146a-5p [CCL5], miR-146b-5p [CCL5], miR-155-5p [WNT5A], miR-15a-5p [ACTR2], miR-16-5p [ACTR2], miR-17-5p [GBP3], miR-20a-5p [GBP3], miR-23a-3p [CCL7, GBP3], miR-23b-3p [CCL7, GBP3], miR-26a-5p [FCAR, GBP1, WNT5A], miR-30e-3p [ACTR2, CCL7,GBP1], miR-32-3p [ACTR2, CD58], miR-374a-5p [ WNT5A], miR-7-5p [FCAR, TLR4] | Cellular response to interferon-gamma |
| 3 | miR-1-3p [PML, VEGFA], miR-146a-5p [NOX4], miR-146b-5p [NOX4], miR-148b-3p [DPP4, TFRC], miR-15a-5p [VEGFA], miR-16-5p [VEGFA], miR-17-5p [CREB1, EGLN3, MMP2, VEGFA], miR-20a-5p [CREB1, EGLN3, MMP2, VEGFA], miR-221-3p [DDIT4, SOD2], miR-221-5p [PML], miR-223-3p [DDIT4, SOD2], miR-23a-3p [EGR1], miR-23b-3p [EGR1], miR-26a-5p [ADM, CREB1, DPP4, PLOD2], miR-29a-3p [DPP4, VEGFA], miR-30e-3p [CREB1, EGR1, SOD2], miR-32-3p [EGLN3], miR-374a-5p [VEGFA], miR-7-5p [ACVRL1, DDIT4, EGLN3, TFRC] | Response to hypoxia |

| | | |
|---|---|---|
| 4 | let-7g-5p [COL4A1, ERO1A, PXDN], miR-148b-3p [COL4A1], miR-15a-5p [B4GALT1, FURIN], miR-16-5p [B4GALT1, FURIN], miR-17-5p [COL4A1, FURIN, MMP2, PXDN], miR-20a-5p [COL4A1, FURIN, MMP2, PXDN], miR-21-5p [MATN2], miR-23a-3p [COL4A1, ERO1A, PXDN], miR-23b-3p [COL4A1, ERO1A, PXDN], miR-26a-5p [B4GALT1, COL5A1], miR-29a-3p [COL15A1, COL4A1, COL5A1, COL5A3, PTX3, PXDN], miR-30e-3p [MATN2], miR-32-3p [BCL3], miR-374a-5p [COL4A1], miR-582-5p [COL5A1], miR-7-5p [MMP19] | Extracellular matrix organization |
| 5 | let-7g-5p [FCAR, IL6], miR-142-3p [PDE4B], miR-148b-3p [ABCA1, PDE4B, SERPINE1], miR-155-5p [CEBPB, WNT5A], miR-15a-5p [CD40, LITAF], miR-16-5p [CD40, LITAF], miR-17-5p [ABCA1, B2M], miR-20a-5p [ABCA1, B2M], miR-23a-3p [PDE4B, TNFAIP3], miR-23b-3p [PDE4B, TNFAIP3], miR-26a-5p [FCAR, PDE4B, WNT5A], miR-29a-3p [TNFAIP3], miR-30e-3p [SERPINE1], miR-32-3p [TNFAIP3], miR-374a-3p [SERPINE1], miR-374a-5p [CEBPB, PDE4B, TNFAIP3, WNT5A], miR-7-5p [FCAR, PDE4B, TLR4] | Cellular response to lipopolysaccharide |
| 6 | let- 7g-5p [C2, TREML1], miR-1-3p [MR1, PML], miR-146a-5p [IFIT3], miR-146b-5p [IFIT3], miR-155-5p [JCHAIN, TLR6], miR-15a-5p [CLU, TREM1, TLR1], miR-16-5p [CLU, TREM1, TLR1], miR-17-5p [B2M], miR-20a-5p [B2M, PCBP2], miR-21-5p [ANXA1,PCBP2], miR-221-3p [IFIT2, PARP9, PRDM1], miR-221-5p [PML], miR-223-3p [IFIH1, LACC1, PRDM1], miR-23a-3p [PRDM1], miR-23b-3p [PRDM1], miR-26a-5p [C1S, PARP14], miR-29a-3p [PTX3], miR-30e-3p [ANXA1, APOBEC3A, PRDM1, TRIM22], miR-32-3p [PEDM1], miR-374a-3p [CLEC5A], miR-374a-5p [ANXA1, CLU, PARP9, PRDM1], miR-582-5p [TRIM22], miR-7-5p [TLR4, TREM1, TRIM58] | Innate immune response |
| 7 | let- 7g-5p [OLR1], miR-146a-5p [CCL5], miR-146b-5p [CCL5], miR-148b-3p [ITGA5], miR-21-5p [OLR1], miR-26a-5p [ITGA5, SELP], miR-29a-3p [F11R], miR-30e-3p [ITGA5], miR-3613-5p [F11R], miR-374a-5p [SELE] | Leukocyte cell-cell adhesion |
| 8 | miR-148b-3p [DDAH1], miR-155-5p [DDAH1, TLR6], miR-15a-5p [CLU], miR-16-5p [CLU], miR-221-3p [SOD2], miR-223-3p [SOD2], miR-29a-3p [PTX3], | Positive regulation of nitric oxide biosynthetic process |

| | |
|---|---|
| | miR-30e-3p [SOD2], miR-374a-5p [CLU], miR-7-5p [TLR4] |

**Table S11. Upregulated miRNAs and their target genes pathways**

| S.NO. | miRNAs and their Targets | KEGG pathways |
|---|---|---|
| 1 | miR-122-5p [CD40LG, TNFSF14], miR-125b-5p [BCL2] miR-134-5p [TNFSF14], miR-215-5p [TRAF5], miR-342-5p [CARD14], miR-629-5p [ZAP70] | NF-kappa B signaling pathway |
| 2 | let-7b-5p [FASLG], miR-122-5p [CD40LG], miR-629-5p [CD28] | Allograft rejection |
| **S.NO.** | **miRNAs and their Targets** | **GO pathways** |
| 1 | let-7b-5p [MAP3K9], miR-125b-5p [CDKL5, MAP3K9], miR-145-3p [PASK], miR-320b [CDKL5], miR-320c [CDKL5], miR-320d [CDKL5], miR-320e [CDKL5], miR-342-5p [EPHA1, MAP4K1], miR-543 [TXK], miR-629-5p [CAMK4, ZAP70] | Protein autophosphorylation |

| | | |
|---|---|---|
| 2 | miR-10a-5p [GATA6], miR-10b-5p [GATA6], miR-125b-5p [ETS1], miR-193a-5p [VEGFB], miR-320b [BTG1], miR-320c [BTG1], miR-320d [BTG1], miR-342-5p [EPHA1], miR-543 [SEMA5A, SIRT1] | Positive regulation of angiogenesis |
| 3 | let-7b-5p [RASGRP1], miR-543 [BANK1, IKZF3], miR-629-5p [ZAP70] | B-cell activation |
| 4 | Let-7b-5p [CCR7], miR-320b [CCR7], miR-320c [CCR7], miR-320d [CCR7], miR-629-5p [CD28, ZAP70] | Negative thymic T cell selection |
| 5 | miR-122-5p [CD40LG, TNFSF14], miR-134-5p [TNFSF14], miR-629-5p [CD24, CD28] | T cell co stimulation |
| 6 | let-7b-5p [ZNF268], miR-125b-5p [DIS3L2, ETS1, FKTN], miR-134-5p [ZNF268], miR-145-3p [ZNF268], miR-193a-5p [RARB], miR-215-5p [GDF11], miR-320b [BTG1, ING5, INSM1], miR-320c [BTG1, ING5, INSM1], miR-320b [BTG1, ING5, INSM1], miR-320d [BTG1, ING5, INSM1], miR-320e [PTPN14, ZNF268], miR-543 [ING5, NPM1], miR-629-5p [BCL11B] | Negative regulation of cell proliferation |
| 7 | let-7b-5p [FASLG, MEIS2], miR-10a-5p [GATA6], miR-10b-5p [GATA6] | Response to growth factor |
| 8 | let-7b-5p [RASGRP1], miR-122-5p [TNFSF14], miR-134-5p [TNFSF14], miR-629-5p [CD28, ZAP70] | T cell activation |
| 9 | let-7b-5p [EDN1, TGFBR3], miR-125b-5p [PLCH2, SH3BP5L], miR-134-5p [ITSN1], miR-145-3p [PASK], miR-193a-5p [DCLK1, ITSN1], miR-342-5p [MAP4K1], miR-543 [AKAP7, DCLK1], miR-629-5p [CAMK4, ZAP70] | Intracellular signal transduction |
| 10 | miR-125b-5p [TOX, BCL2] | CD8-positive alpha-beta T cell lineage commitment |
| 11 | miR-122-5p [CUBN, FKBP5], miR-125b-5p [NFIB], miR-320b [BNIP3], miR-320c [BNIP3], miR-320d [BNIP3], miR-543 [BANK1, IKZF3, SLC11A1] | Response to bacterium |
| 12 | let-7b-5p [TGFBR3], miR-122-5p [FMNL3, PTPRB], miR-125b-5p [FMNL3], miR-193a-5p [VEGFB], miR-320b [PTPRB], miR-320c [PTPRB], miR-320d [PTPRB], miR-342-5p [EPHA1], miR-543 [PTPRB, SIRT1, TGFBI] | Angiogenesis |

| | | |
|---|---|---|
| 13 | miR-122-5p [ADAM33, CD40LG], miR-543 [ADAM23, TXK, ITGA9], miR-145-3p [ADAM23] | Integrin-mediated signaling pathway |
